# Social disparities in triple-negative breast cancer incidence and severity at diagnosis in Greater Paris, France: confronting race and ethnic blindness

**DOI:** 10.1101/2025.07.10.25331284

**Authors:** Pierre Chauvin, Thomas Huet, Joseph Gligorov, Carolyn Sargent

**Affiliations:** Department of Social Epidemiology, Institut Pierre Louis d’Epidémiologie et de Santé Publique, Sorbonne Université, INSERM, Paris, France; French Institute for Demographic Studies (INED), Aubervilliers, France; Department of Oncology, Hôpital Tenon, Assistance Publique-Hôpitaux de Paris, Sorbonne Université, Paris, France; Department of Anthropology, Washington University in St. Louis, St. Louis, United States

## Abstract

Triple-negative breast cancer (TNBC), an aggressive subtype with poor prognosis, is of higher frequency in African-American women and women of Sub-Saharan African origin. In France, legal constraints on obtaining health data on race, ethnicity, or nationality in cancer registries and medical records make it difficult to estimate the prevalence of TNBC according to women’s origins. These constraints result from a historical “universalist” approach to French citizenship which prohibits the routine collection of ethno-racial data. An anonymous, statistical survey we conducted from the medical records of 780 women with breast cancer followed in a university hospital in Paris showed that TNBCs were at least 3 times more common in women born in Sub-Saharan Africa than in women born in France. The former consulted at a more advanced stage of the disease than the latter. The results of an ethnographic study of African women in the Paris region with breast cancer, conducted for several years, highlighted some explanatory factors: low breast cancer awareness, perceived causes far from biomedical etiology, the weight of shame and secrecy, prior recourse to local healing, difficulties in communicating with health professionals and navigating the healthcare system. Considered a public health priority, TNBCs are an emblematic example of the limits produced by French race and ethnicity blindness in public health, epidemiology, prevention and health care.

## Introduction

Triple-negative breast cancers (TNBC), characterized by the absence of receptors for estrogen, progesterone and the HER2 protein, form a subtype of breast cancer that is usually more aggressive than the others, responding less well to available treatments, and with poorer prognosis [1]. In mainland France, it is the least common subtype of breast cancer but highly significant in clinical and social terms: it represents 15% of cases and mainly concerns women under 40, pre-menopausal, too young to be part of the target population of the national organized screening program (which in France concerns women aged 50 to 75) and tested at an often advanced stage [2,3]. Accordingly, in this supplement to the scholarly literature, ethno-racial differences in TNBC prevalence as well as cross-cultural questions of how breast cancer is distributed within and among social communities merit further examination.

This article presents epidemiological and social anthropological research findings on ethno-racial disparities in the causes and distribution of TNBC in particular populations, with special attention to France. We explore TNBC among Sub-Saharan immigrants in France and their apparent vulnerability to this high-risk form of breast cancer. We frame this analysis in the context of contested constructs of race and ethnicity in various countries and how these play out in the recognition of breast cancer risk in certain populations. France is a unique case to center our discussion because of the historical and contemporary dissension about race as an individual and collective category of identification. We will discuss state regulations regarding data collection on race and ethnicity and the implications of these policies for understanding breast cancer risk, diagnosis, and treatment.

### Ethno-racial disparities in TNBC

To better understand the French case, we provide comparative data from the US, UK, and Africa, focusing on racial and ethnic inequalities in TNBC prevalence in these regions.

### Ethno-racial disparities of TNBC in the United States

In the United States (US), racial inequalities in breast cancer incidence and mortality have been known and described for decades [4]. Mortality has consistently been observed to be higher among African-American women than among non-Hispanic White women, while differences in incidence are smaller (and age-dependent; breast cancer being more common among African-American women under 35 years old than among non-Hispanic White women of the same age, the opposite being observed after 40 years old). Initially, the higher mortality and shorter survival of African-Americans had been attributed to their lower socioeconomic status and their difficulties in accessing diagnosis and then care, resulting in a late stage at diagnosis [5]. However, in many studies, these racial differences persisted after adjustment for socioeconomic characteristics, access to care and stage at diagnosis, suggesting a possible difference in the nature of the disease itself, in terms of aggressiveness. Indeed, since the 1990s [6], many studies have accumulated the evidence of racial disparities in terms of presence or absence of one or more tumor markers [7]. In 2020, a review of the literature reported the details of the prevalence of TNBC by race from some thirty US studies (published mainly between 2007 and 2014) to conclude that it is twice as high among African-American women as among non-Hispanic Whites in all age groups [8]. Although BRCA1 gene mutations are a recognized genetic risk factor for TNBC, their prevalence in African-American women with breast cancer is of the same magnitude, or even lower, than that in non-Hispanic (and non-Ashkenazi Jewish) white patients: approximately 1% or 2% [9,10]. Other genetic abnormalities associated with a high risk of TNBC have been discovered more frequently in African Americans [11,12,13]. It is worth noting that US research most often, if not always, begins from ethno-racial categories, only then (and rarely) to question the role of geographical origins. For example, it is within the population of African-American women with breast cancer (identified in the 1998-2015 databases of the National Program of Cancer Registries and the US Cancer Statistics which cover 99% of the US population) that Sung et al. studied the disparities in TNBC prevalence according to the country of birth [14]. They were significantly lower among African-American women born in the Caribbean (21.2%) or in East Africa (11.6%) than among African-American women born in the US (23.7%). The difference was not significant between the latter and Black women born in West Africa (24.1%). The authors emphasize that “*nativity and geography origin in black women has rarely been examined, although considering nativity-related differences may improve our understanding of the etiologic heterogeneity of breast cancer*”. In the end, their results plead for a genetic base of racial disparities in TNBC since their frequency seemed proportional to the importance of the shared genetic ancestry among the 4 groups compared.

After an initial diagnosis, in the population of women with TNBC, African-American women have a 5-year survival that is 2.5 times shorter than white American women (14% versus 36%) and an excess mortality of around 30% [15]. Many reasons can explain these racial differences in prognosis within TNBC patients in the US context: higher prevalence of obesity and other comorbidities, fewer cancer screening opportunities, later diagnosis, barriers in access to cancer care in the population of African-American women [16]. However, these differences remained pronounced after adjustment for all these factors, strongly suggesting, here too, a biological origin [17,18]. In fact, it has been shown that at the cellular level, TNBC tumors in African-American women were more aggressive than those in White women and numerous differences in gene expression and molecular phenotype have been observed between these two groups [19]. In a perspective on the genomics of human stress, some authors consider these biological (acquired) differences as the result of a cascade of macro, meso and micro social determinants (fairly usual in the conceptual models used in social epidemiology) of social stress, having ultimately a molecular impact down to the cellular level [20].

### Ethno-racial disparities of TNBC in the United Kingdom

In England, in a study limited to the North East London Cancer Network in 2005-2007 [21], the frequency of TNBC was 8% in White women, 19% in South Asians (TNBC also being more common in the Indian subcontinent [22,23]) and 25% among Blacks. Such differences were also observed (even if lower) in a prospective hospital cohort of women with breast cancer aged under 40 years included between 2000 and 2008 in multiple sites in the United Kingdom (UK): 18.6% of cases among Whites versus 28.6% of cases in Black women were TNBC [24]. The English National Cancer Registry has been collecting individuals’ ethnicity with a usable completion rate (≥ 90%) since 2013. The exhaustive study of racial disparities in breast cancers is therefore quite recent (published in 2021, on 2013-2018 data) [25]. Unfortunately, its authors did not report the frequency of TNBC (perhaps because of the frequency of missing data concerning these 3 markers) but estimated that “Black African” patients were more at risk of late-stage, high-grade and/or ER negative tumors than Whites at all ages at diagnosis. A lower but significant risk was also highlighted among “Black Caribbean” women aged 53 to 70 at the time of diagnosis.

Another recent study, based on the same English national registry, observed a higher risk of diagnosis at a late stage of breast cancer in “African” women and, to a lesser extent, in “Caribbean” women compared to White women (including after adjustment for year of diagnosis, age, comorbidities, deprivation index of their area of residence and region) [26]. Interestingly for a European perspective, in the context of a universal national health service and the existence of an organized breast cancer screening program, the authors put forward several possible explanations for the differences observed (in addition to the TNBC ethno-racial differences and their greater aggressiveness). Indeed, as observed in some quantitative British studies, faced with cancer, women from minorities, particularly Black women, may recognize fewer cancer symptoms [27,28] and manifest higher cancer fatalism [29], resulting in delayed use of primary health care [30].

### TNBC in North Africa and Sub-Saharan Africa

In Africa, a literature review and meta-analysis of 67 publications on TNBC prevalence was published in 2018 [31]. In most countries except in North Africa, survey samples are small (a few hundred cases at best) and most often focus on the very small minority of breast cancer cases who have access to tertiary health care. In the absence of high-quality epidemiological data (population-based cancer registries only exist in 20 of the 46 African countries and are of very variable quality [32], particularly for the analysis of tumor markers [33]), these estimates provide only orders of magnitude. Estimates showed that TNBC was by far the most frequent in West Africa (45.7%, 95% CI = [38.8%-52.8%]), comparing with East Africa (25.0%), Maghreb (i.e. Morocco, Algeria and Tunisia in Northern Africa, 23.0%), Southern Africa (20.4%) and Central Africa (14.8%) %) [34].The difference in TNBC prevalence between West and East Africa was even greater in a study which compared Ghana (53%) and Ethiopia (9%) respectively [35]. Data has only been published in 6 of the 14 French-speaking West African countries (the countries of origin of the vast majority of Sub-Saharan African immigrants in France). In Mali (n=114) [36], Sénégal (n=522) [37], Côte d’Ivoire (n=335) [38], Togo (n=117) [39], Bénin (n=244) [40], and Guinea (n=56) [41], respectively 46%, 47%, 43%, 38%, 32% and 26% of women consulting the capital’s referral hospital for breast cancer were TNBC cases. Estimates of BRCA1 mutation prevalence in Sub-Saharan Africa remain sparse and based on small sample sizes [42]. For instance, in a cohort of 434 unselected Nigerian women with breast cancer, 7% had a BRCA1 mutation [43]; this means that it cannot explain the very high prevalence of TNBC (estimated between 12.2% and up to 49.4% according to different regional studies [44,45].

### Questioning race/ethnicity in France

The way in which race/ethnicity statistics are defined, collected, and used in social and health sciences varies considerably between France and the US, Canada and the UK. In these last three countries, ethno-racial categories have been used routinely for decades even if they have changed over time and their relevance is regularly questioned [46,47]. The UK is the only country in Western Europe to interrogate ethnicity in the general census of its population (since 1991) and to produce routine ethnic statistics (notably health statistics) [48].

### Ethno-racial characterization: between rejection by principle and popular uses

Contrary to widespread belief, France is far from being the only European country not to do so, even if the issue of race/ethnicity is particularly sensitive here [49]. Indeed, the criminal use of the notion of “race” during the Second World War by the Nazi regime in occupied Europe meant that the notion of race was completely abandoned in all the countries of Western continental Europe after the Second World War. In 1952, Claude Lévi-Strauss wrote that “*the original sin of anthropology consists of the confusion between the purely biological notion of race (assuming that […] this notion can claim objectivity) and sociological and psychological productions of human cultures*” [50]. In a 1982 book, Albert Jacquard (a celebrated and popular scientist in biology and genetics in France) asserted that, for modern genetics, the notion of race was absurd since the majority of genetic variability is within geographical groups, and not between them [51]. We only quote these two scientists briefly to illustrate to what extent, in France, the notion of race and ethnicity has been historically and epistemologically discussed in terms of socially constructed groups (some French social scientists speak of “racialized” groups) and widely discredited as a biological attribute in medicine and epidemiology (with the notable exception of hemoglobinopathies and G6PD deficiencies [52]).

Contrary to the US, where history, marked by slavery, racial segregation and struggles for civil rights, has led to an institutionalization of racial categorization, France has historically had a “universalist” approach to citizenship, where the emphasis is on national unity rather than racial or ethnic differences (for a much more detailed comparison between the US and France on their visions of race and origins, respectively, French-speaking readers can refer to a reference article by F. Héran [53]). Apart from its colonies (where racial statistics were in common use, associated with extreme racist prejudices [54]), the French Republic has adopted an assimilationist paradigm that favors the integration of immigrants and their descendants without explicit recognition of ethno-racial identities [55]. The first article of the French Constitution stipulates that France “*ensures the equality of all citizens without distinction of origin, race or religion*” and, therefore, prohibits any counting of these categories of population. This principle is of course debatable and widely discussed in public and political debate and by French researchers, particularly in the social sciences and in the field of studies on discrimination. For example, P. Simon speaks of the “*choice of ignorance*” and criticizes this posture of “*equality through invisibility*“ [56] and F. Héran calls for “*stopping opposing republican principles to ethnic statistics*” [57]. Also in public health and social epidemiology, French researchers regularly regret that ethno-racial characteristics cannot be easily queried, e.g. recently during the Covid-19 epidemic which particularly affected areas with a high proportion of immigrants [58,59]. Thus, if the assimilation of immigrants in France is an objective reality (as evidenced by a growing diversity of marriages or partnerships at each generation that is infinitely more frequent in France than in other Western countries [60]), at the same time experiences of discrimination are 3 to 4 times more frequent among immigrants or children of immigrants of Caribbean, African or North African origin [61]. In common language, the words “race” and “ethnic group” are certainly never used in France but categories of “origin” are widely attributed and used (either objectively, according to people’s ancestry, or subjectively according to physical appearances and/or cultural prejudices): In France it is common to speak of “Arabs”, “Africans”, “Caribbeans”, “Asians”, etc. In popular parlance, the words “*blanc*”, “*toubab*” (derived from the Arabic word “*toubib*” [doctor] and used in the broader sense of “White” in French-speaking West African countries), “*black*”, “*noir*”, “*rebeux*” (a backwards slang word for Arabs), “*jaune*” or “*Chinois*” (whatever their real origin) and so forth refer directly to the skin color of people. A large, multipurpose, demographic survey dedicated to French population’s origins conducted in 2008 showed that class dimensions were stronger than ethno-racial ones in shaping self-perceived personal identity in France but suggested also that blackness was a crucial dimension of ethno-racial identification [62].

### The legal background to collect data on “origins”

There is a general principle in French legislation which prohibits the collection of data on race or ethnicity in public and private statistics. But, in 2007, the French Constitutional Court reminded that the processing of personal data that directly or indirectly reveal people’s racial and ethnic origin, and the introduction of race or religion variables in administrative files, to be contrary to the French constitution, as well as the a priori definition of an ethno-racial reference system. But, at the same time, it indicated that *“the processing necessary for carrying out <u>studies</u> on measuring the diversity of people’s origins, discrimination and integration can relate to objective data on people’s ancestry*”. In the commentary to its decision, it explained that *“these objective data may, for example, be based on the name, geographical origin or nationality prior to French nationality.”* In addition to these “*objective data*”, it authorizes the collection of data based on the *“feeling of belonging”* to an ethnic group in specific surveys. So, for research purposes only, these data can be collected with special authorization which must be argued and justified regarding the scientific objectives pursued. They are considered as “*sensitive data*”, in the same way as political opinions, religious affiliation or sexual orientation, whose collection can only be exceptional and strictly regulated by the French data protection authority. At no time did the Constitutional Court open the possibility of <u>routine</u> statistics according to ethno-racial categories but allows the detailed collection of nationalities and countries of birth over several generations in specific surveys. Although such statistical studies on detailed origins have been emerging in France over the past decade, particularly in demography and sociology, they remain rare in epidemiology and almost non-existent in clinical research. Patients’ origin may possibly (but rarely) be mentioned in their medical record when their life circumstances are briefly detailed but no doctor would systematically categorize their patients according to their race or ethnicity.

### The diverse origin of the French population

In the absence of ethno-racial statistics, only estimates from unreliable sources circulate on the proportion of Blacks in France: between 3.5% and 6.5% (French overseas regions (FOR) included) [63], i.e. approximately the same proportion as “Blacks” (1.98%) and “Indians” (2.30%) altogether in the UK, and 2 to 4 times less than in the US (where 12.4% of the population is “African-American”). Another estimate (just as poorly sourced but plausible [64]) postulated in 2004 that 3.5% of the French mainland population was Black, broken down into 25% from the FOR (including the two Caribbean islands in the French West Indies), and 75% of Sub-Saharan African origin (most of whom having immigrated to France since the 1980s [65,66]).

In the Paris region in 2008, 43% of the population aged 18 to 50 had a direct link with immigration to mainland France over two generations, in the sense of being immigrants or descendants of immigrants, or natives from FOR or descendants from FOR: 11% are of European origin, 14% from the Maghreb, 7% from Sub-Saharan Africa, 4% from FOR, 3% from Asia, 1% from Turkey, and 3% from other regions [67]; which possibly leads to a (very rough) estimate of 11% of “Blacks” in the Paris regional population. In France, strictly speaking, in the general population census and public statistics, “immigrants” are people living in France who were born abroad and who are not French citizens by birth. Whether or not they were subsequently naturalized as French citizens, they are still counted as immigrants. For their part, French people born abroad are not immigrants, nor are French people born in French overseas regions (such as the French Antilles) who have settled in mainland France. So, if we only count immigrants, 17.9% of the population of the Paris region are immigrant: half of which (48.3%) from Africa: 29.7% from the Maghreb and 18.6% from Sub-Saharan Africa respectively. Four countries of origin account for half of the latter: Mali (2.6%), Côte d’Ivoire (2.2%), Senegal (2.1%) and Congo (2.0%).

### Triple negative breast cancers: the case of the French West Indies

In the absence of ethno-racial statistics available in France, it is interesting to look at the epidemiology of breast cancer in the French West Indies, two French overseas islands where black people are the overwhelming majority of the population. It is composed of descendants of African slaves (and more marginally of Indian forced laborers who succeeded the slaves on the plantations after the abolition of slavery in 1848) and white settlers, as well as metropolitan expatriates in the modern era. Both islands (Guadeloupe and Martinique) have a cancer registry.

In Martinique particularly, society remains very marked by socio-racial stratification which dates back to the era of slavery: “*the stereotypes and prejudices attached to skin color and racial categories, which are in reality emic categories, are always extremely operational*” [68]. The epidemiology of TNBC in this island of 367,000 inhabitants has only recently been published [69]: the proportion of TNBC is 20.9% (compared to 15% in mainland France) with a 5-year survival rate of 70.1% and a 1-year survival rate of only 34.1% for metastatic forms. In this paper, the race issue is furtively raised at the beginning of the discussion in a single sentence (“*TNBC is a subtype with a higher prevalence in women younger than 40 years old with African or Asian ancestry or non-Hispanic Black or Hispanic women, as is the case in Caribbean populations*”), then is implicit when comparing TNBC among other Caribbean islands…Not only are ethno-racial statistics not provided (since this is impossible) but the racial question is never addressed in the conclusive recommendations on the identification and management of TNBC (perhaps precisely because the racial question remains sensitive on this island). On the other hand, in 2017, in the only article on TNBC in Guadeloupe (the other French Caribbean Island, with a population of 379,000) where TNBC accounted for 14% of all cases, and were more frequent in patients under 40 (21.6% vs. 13.4%, p=0.02), the authors not only cite a substantial bibliography on TNBC in African-American women in the US but also explicitly outline the “*lack of data on ethnicity and socio-economic status*” in French cancer registries as the main limitation of their study [70].

### The absence of ethno-racial data on cancer in France

In the absence of systematic collection of patients’ race or ethnicity, and considering the absence of details on their origin (i.e. their nationality and country of birth, and those of their parents) in any of the existing routine health information systems, it is impossible in France to easily study the biological and clinical disparities of diseases according to one or the other of these two characteristics. Notably, the 25 existing French Cancer Registries collect neither of them, nor any socio-economic characteristics of people (for example, there is no data on educational level or occupation). Social inequalities in cancer incidence are only understood indirectly, through ecological analyses which use socioeconomic statistics of people’s area of residence [71].

On the other hand, in computerized hospital administrative files (in outpatient consultation as well as in hospitalization), only patients’ gender, date and country of birth are collected systematically, along with their address and health insurance status. These files can be reconciled with medical records even if this has to be done manually.

### Both the US ethno-racial and the French classification of origins have their own limits for the study of TNBC

Not only do these two classification systems make it difficult to compare inequalities in the prevalence of TNBC, but each has its own limitations.

In the US, from a biological perspective, the category “African American” actually tells us little about the ancestry of this population: genome-wide ancestry estimates of self-reported African Americans show an average proportion of 73.2% African, with notable differences across states in the US (between 64% and 81%) [72]. It would still be necessary to know which African origins we are talking about, since we saw previously [33] that the prevalence of TNBC varies considerably today between West, East, Central, and South Africa.

In France, the collection of French nationality (at birth or acquired subsequently) and country of birth in the population census makes it possible to distinguish French nationals from immigrants according to their country of birth (but not systematically in health information systems, nor in epidemiology as mentioned above). In 2025, the introduction of the parents’ place of birth (in France versus abroad, but without specifying in which country) in the population census will make it possible to distinguish French nationals born to immigrant parents. More detailed demographic surveys (the TeO surveys being a reference in France [73]) or some rare epidemiological surveys [74,75] have also collected the nationality and country of origin of parents, with special permission.

As we can see, the collection of “origins” can be more or less detailed. Given the complexity of migratory and historical links due to colonization (and also of the changing rules for acquiring French nationality, by birth or descent, over time), misclassification errors are not rare. For example, people born French abroad to French parents can be either “white” settlers or “natives”. Even in its most detailed form, this characterization tells us little (and probably even less than the racial categorization of the US) about the ancestry of people.

### Objectives

By means of data from two research projects, one archival and one ethnographic on Sub-Saharan immigrant women living in France and undergoing treatment for breast cancer, we assess the potential etiological factors (genetic, environmental, political, economic) associated with TNBC, as we noted, a particularly aggressive and challenging form of cancer. This paper therefore presents an original perspective on ethno-racial disparities found in epidemiological studies of TNBC; statistical analyses of medical records on breast cancer patients; and ethnographic research on the lived experiences of breast cancer among Sub-Saharan women living in France.

## Material and methods

### A quantitative analysis based on patients records in a Paris University Hospital

Three samples of patients were selected from the total number of patients with breast cancer followed in the oncology department of Tenon Hospital (a university hospital located in the east of Paris, in a socially diverse, mainly working-class, neighborhood) between 01/01/2000 and 12/31/2017: 1) all the 180 patients born in Sub-Saharan Africa (SSA); 2) a random sample of 200 patients born in the Maghreb (Morocco, Algeria, Tunisia: MAG); and 3) a random control sample of 400 patients born in France (FRA). Indeed only the country of birth was available in patients’ files.

Let us emphasize from the outset the limits of this proxy for characterizing the ethno-racial origin of women as it shows possible classification bias. Of course, a “White European” woman may be born in Africa (for example to French expatriate parents), vice versa (and surely more often) a “Black” woman can be born in France (for example to Black African immigrants, to French Caribbean parents, or with an even more distant black ancestry). Furthermore, by limiting ourselves to women born in France as a proxy for “White European women”, we excluded from our sample all those who were born in another EU country (in particular in Southern EU, which account for a significant – although aging – proportion of the immigrant population in France).

Initially, the required sample size was calculated based on our primary hypothesis regarding the difference in TNBC prevalence between SSA (p1) and FRA (p2), with the following assumptions: p1=30%, p2=15%, with α=0.05 (two-sided test) and 1-β=0.90. At least 159 subjects per group had to be included. For greater power, we chose to include twice as many FRA as SSA. Finally, we chose to add a third (exploratory) group of women born in North Africa (MAG).

The study period covering a time when medical files were not or only minimally computerized, all of the paper files of the selected patients were retrieved from the department’s archives under the supervision of the department’s head oncologist, then anonymized, re-read and manually analyzed one by one. After excluding empty files and those concerning patients who only occasionally consulted for a second medical opinion but who were not followed in the department, the numbers of the 3 populations studied were 177 SSA, 153 MAG and 336 FRA women.

For each patient, the following characteristics were collected anonymously from their paper medical record: age, health insurance status, professional situation, level of deprivation of their neighborhood of residence, results (positive or negative) of ER, PR and HER2 tumor markers and its TNM stage (the TNM staging system standing for tumor, node and metastasis). Health insurance status was coded into 4 modalities: usual health insurance by Social Security (SS); specific health insurance for low income people (designated by the acronym CMUc for Universal Supplementary Health Coverage, it is granted below a certain threshold of household income: for example €810 for a single person); specific health insurance for the undocumented (designated by the acronym AME for State Medical Aid, also subject to a resource ceiling and accessible after 3 months of presence in France); and paid care. This last category concerns (non-EU) foreigners who do not live in France but can afford to travel to seek treatment in France: their care is then fully billed to them by the hospital (but may be covered in their country of origin by their own public or private health insurance). As we can see, patients’ insurance status gives several indications on the patients’ situation: poor, in an illegal situation, or foreign citizens living abroad.

The level of deprivation of the neighborhood of residence used the FDep [76] (for French Deprivation index), a composite indicator used in France since 1990 and updated at each population census. It was created to provide a geographic aggregate indicator of social disadvantage specifically adapted to health studies on the French population, when individual socioeconomic data are not available. It is calculated from the unemployment rate, the proportion of blue collars, the proportion of baccalaureate holders and the household median income per consumption unit in the area. Here, we used the FDep calculated in 2015, at the finest geographic level routinely available in France: the “neighborhood of residence” (or IRIS, an acronym for *smallest spatial area for statistics*). Indeed, in France, IRIS are the finest spatial divisions for which aggregated population census data are available. Their average population is 2000 inhabitants in the Greater Paris area, for an average surface area of 0.42 km^2^ (i.e. a circle of 370 meters radius). They are homogeneous in terms of the type of housing and their limits are based on major cuts in the urban fabric (main roads, railways, waterways, etc.). The distribution of the FDep is conventionally divided into quintiles (the first quintile corresponding to the most advantaged neighborhoods). Each patient’s neighborhood of residence was identified based on the geocoding of their full address. Among women born in France, only 3 addresses (out of 336) were not found. Among women born in SSA, 22% came from abroad to seek cancer care at Tenon Hospital. Most declared an accommodation address with a relative in France (this accommodation address was geocoded), except 9 (out of 177) for whom no accommodation address was reported. Among women born in the Maghreb, 13% came from abroad and all except 6 declared an accommodation address with a relative in France (here too, it is this accommodation address, which was geocoded, and we therefore have 6 missing data on the 153 women concerned).

The data were collected between 06/29/2016 and 08/03/2018. French law does not consider the retrospective reuse of existing data (e.g., from archived medical records) as “research involving human beings” and therefore does not require approval from a national ethics committee. Moreover, at that time, retrospective studies conducted at the Paris University Hospitals did not require special authorization if they were conducted in a single care department, under the supervision and responsibility of its department head, a university professor (here, the third author of this article), and 2) if no nominative data was collected and used by researchers outside the department’s medical team. At the time the paper medical records were compiled, the retrospective reuse of anonymous data did not require formal patient consent.

Proportions between subgroups were compared using the Chi-square test and age means using the two-sided Student t-test. For multivariate analysis, two logistic regression models were fitted, estimating adjusted Odds ratios (OR) and their 95% confidence intervals (CI) All statistical analyses were performed using IBM^®^ SPSS^®^ Statistics software v. 24.0 (Armonk, NY: IBM Corp, 2016). To assess multicollinearity among the explanatory variables, the Variance Inflation Factor (VIF) was calculated for each predictor. Because the proportions of missing data regarding triple-negative status were significantly different in the 3 patient samples (SSA, MAG, and FRA), a sensitivity analysis was performed, assuming that all missing values were positive or negative, respectively.

### An ethnographic study on West African immigrants with breast cancer in the Paris region

To supplement the quantitative and archival research reported above, we discuss a five-year ethnographic study of cancer and sociality among West African immigrants residing in Paris and in treatment for breast cancer. This research on meanings and strategies about breast cancer among immigrants from the Senegal River Valley (Mali, Mauritania, Senegal) and northern regions of Guinea and Cote d’Ivoire living in greater Paris, France, focused on three central questions: 1) how sufferers, families, interpreters and clinicians collectively and progressively define and manage cancer; 2) how social relations are restructured around the collective experience of breast cancer; 3) how transnational meanings and strategies are continuously generated by means of cell phone and personal courier communications between those residing in Paris and in West Africa.

Its methodology has been described in detail previously [77,78]. Briefly, the research team consisted of two anthropologists: the last author of the present paper, and another (see Acknowledgements), a retired midwife trained in ethnographic methods who had established three family planning centers in northern suburbs of Paris where she also offered prenatal and postnatal care, and sexual education (her career was largely devoted to the population on whom we focused in this ethnographic study). Both researchers had previous experience working in West Africa (one of them spoke Wolof and the other Baatonu and Bambara). The team was completed by two interpreters (in Wolof, Bambara and Soninké).

The core of the ethnographic research in this study was the McGill Illness Narrative Interview (MINI) [79]. This questionnaire, translated in French in 2013, is a semi-structured interview schedule that systematically presents questions for patients regarding the history and ongoing process of their health condition. The questions encompass description of the illness symptoms, types of consultations to obtain diagnosis and therapeutic interventions, as well as prognosis, impact on everyday life, and spiritual concerns (see Supporting Information 1). It guided us in framing questions concerning how African women with breast cancer living in the Greater Paris area progressively define, understand, and manage cancer whether using local West African concepts or biomedical vocabulary. The MINI allows replication of questions systematically across patient populations using appropriate terminology (see Supporting Information for the English version of the MINI). Data were interpreted by thematic analysis, a means of identifying patterns in interview transcripts and open-ended surveys, for the purpose of defining and naming codes and themes. The resulting collection of coded themes then constitute the basis for conclusions to the diverse questions to which informants replied.

During the 5-year project, ethnological data on breast cancer in African women data were collected between 01/06/2014 and 31/05/2019 in 7 different ways: 1) we interviewed 36 women in treatment for breast cancer who gave their oral consent, in the presence of their doctor - born in the Senegal Valley River countries (Mali, Sénégal, Mauritania), as well as Guinée and Côte d’Ivoire - at four public hospitals in Greater Paris (Tenon, where the quantitative study was conducted subsequently, Avicenne, Delafontaine, and Saint-Louis), as well as caretakers, clinicians and interpreters; 2) regular home visits were conducted between July 2014 and July 2019 for 10 women to assess kinship obligations, gender hierarchies, and moral conventions, using semi-structured interviews; 3) participant observation in four women’s immigrant associations in northern Paris suburbs allowed us to assess popular representations of breast cancer and treatment modalities; 4) we held focus group discussions on breast cancer with group leaders and members of 50 immigrant women’s associations; 5) the research team also observed oncologist-patient consultations two days weekly for two years; 6) it also accompanied patients from their home to the hospital, visited frequently (sometimes daily) hospitalized patients, and assisted them with access to social services; and 7) we held a meeting with five interpreters of Senegal River Valley languages (Bambara, Soninke, Pulaar, Wolof, Hassania) from *ISM Interprétariat* - a non-profit association which is the main interpreter resource used in Parisian public hospitals - to have a semi-structured discussion about occasions on which these interpreters had been asked to intervene to convey information to patients from Senegal, Mali, Mauritania, or Guinea, whether in person or by telephone.

The ethnographic results presented here are a part of a larger research project on “The Influence of Sociality in Cancer Decision Making”, funded by the National Science Foundation (grant no. 1354336). Verbal informed consent was obtained from all participants in accordance with institutional ethical guidelines. Prior to participation, the study’s objectives and procedures were explained in clear and comprehensible language. Participants provided an explicit verbal agreement to participate, which was documented in the study records, including the date and time of consent. No witnesses were present during the consent process. All data were recorded anonymously to ensure participant confidentiality. The study protocol and consent procedure were reviewed and approved by the Ethics committee at Washington University in Saint Louis (IRB no. 202403075, 18 April 2014).

## Results of the quantitative analysis based on patient records

### Sociodemographic characteristics

The 3 groups of patients differed significantly in terms of age (the SSA being 10 years younger on average than the FRA), poverty rate (45.7% of the SSA benefited from CMUc or AME, compared to 22.3% of MAGs and only 0.6% of FRA) and type of neighborhood of residence (SSA and MAG living 2.5 times more often in the most disadvantaged neighborhoods than FRA: respectively 26.8 %, 29.3% and 11.4%, p<0.001). Regarding their professional situation, there were 2 to 3 times more managers or higher intellectual professions among the FRA (17.9%) than among the SSA (7.3%) or MAG (5.9%) and twice as many more intermediate professions as well. Half of the SSA and MAG had no occupation compared to a third of the FRA. (Table 1).

**Table 1.** Sociodemographic characteristics.

|  | SS Africa<br>n=177 | Maghreb<br>n=153 | France<br>n = 336 |  |
| --- | --- | --- | --- | --- |
| Age: average (sd) | 54.1 (11.9) | 60.7 (12.7) | 65.2 (12.9) | p<0.001 |
| Health insurance status |  |  |  |  |
| SS | 34.5% | 66.0% | 99.1% |  |
| CMUc | 9.0% | 9.2% | 0.6% |  |
| AME | 36.7% | 13.1% | 0.0% |  |
| Paid care | 19.8% | 11.8% | 0.3% | p<0.001 |
| Occupation |  |  |  |  |
| Not in the labor force | 50.3% | 50.3% | 33.3% |  |
| Retirement | 7.9% | 9.2% | 15.2% |  |
| Blue collar | 8.5% | 12.4% | 4.2% |  |
| Lower white collar | 11.9% | 9.2% | 12.5% |  |
| Small business | 6.2% | 4.6% | 2.1% |  |
| Middle white collar | 7.9% | 8.5% | 17.9% |  |
| Higher white collar | 7.3% | 5.9% | 14.9% | p<0.001 |
| Neighborhood deprivation quintile |  |  |  |  |
| 1 (most favored) | 15.8% | 12.4% | 24.7% |  |
| 2 | 14.1% | 19.0% | 22.6% |  |
| 3 | 17.5% | 15.7% | 22.9% |  |
| 4 | 22.0% | 20.9% | 17.6% |  |
| 5 (most disadvantaged) | 25.4% | 28.1% | 11.3% |  |
| unknown | 5.1% | 3.9% | 0.9% | p<0.001 |

### Frequency of TNBC and associated factors

TNBC were more than 3 times more frequent among SSA than among FRA (Fig 1): respectively 38.2% (95% CI = [31.2%-5.8%]), and 12.2% (95% CI = [8.9%-12.2%]). The difference between MAG and FRA was not statistically significant. Note that the results of tumor markers were missing for 5.6% of SSA, 9.2% of MAG and 0.9% of FRA (p<0.01). Sensitivity analysis showed that these differences in the number of missing data cannot explain the discrepancies noted (see Supporting information 2, Table S1).

**Fig 1.**
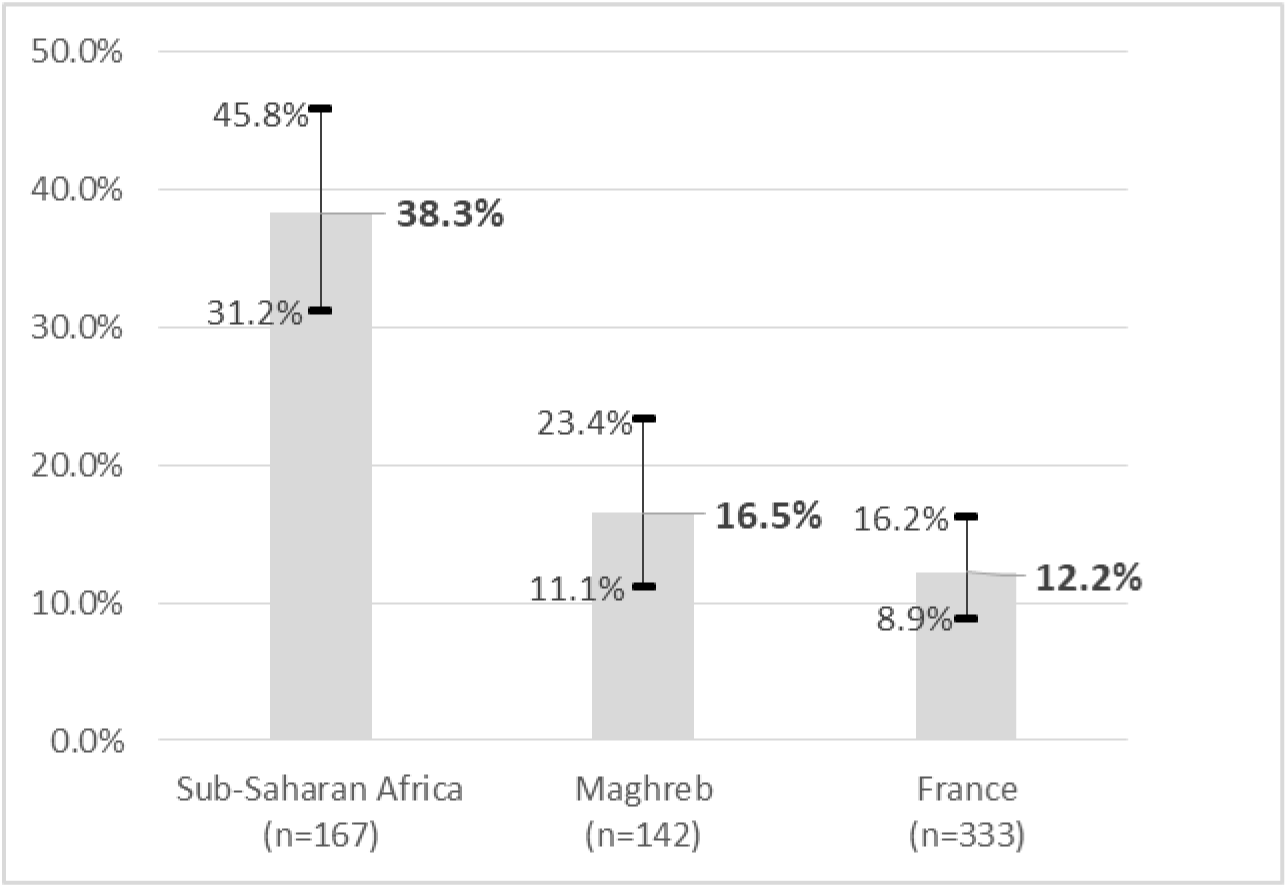
Frequency of TNBC by country of birth (% and 95% CI).

Half of the TNBC were found in SSA and half of the non-TNBC in FRA (Table 2). Women with TNBC were also significantly younger (by 5 years on average), more often undocumented migrants (25.8% versus 9.1%) and/or poor (33.1% versus 13.2%), more often with a low-skilled occupation or not in the labor force (65.3% versus 49.2%, p=0.01). They also lived more often in a disadvantaged neighborhood of Greater Paris (50.0% versus 37.5, p=0.013).

**Table 2.**
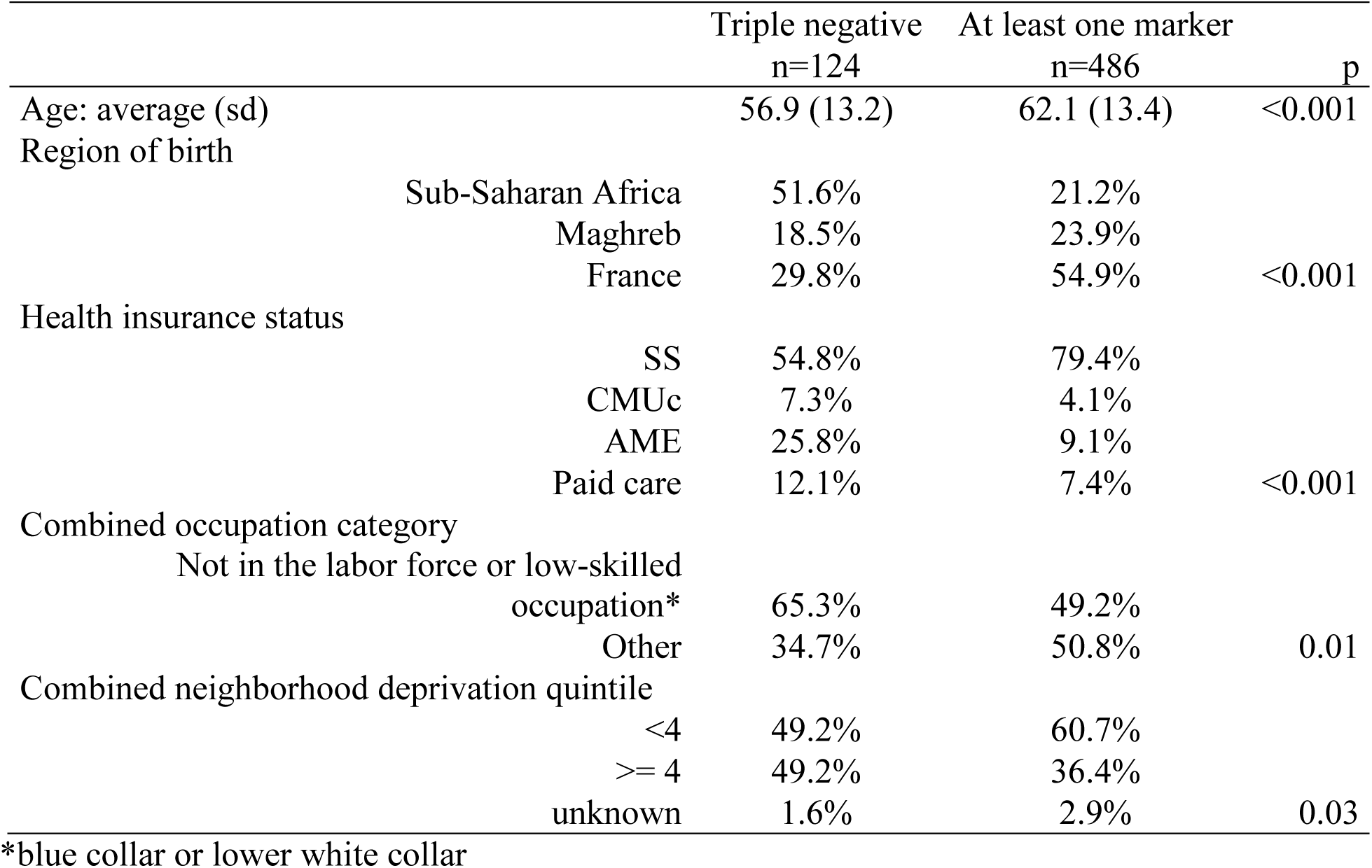
Description of TNBC.

In multivariate analysis, the 3 factors associated with TNBC were being born in Sub-Saharan Africa, being an undocumented migrant in France and/or being inactive or with a low-skilled occupation (Table 3).

**Table 3.** Factors associated with TNBC (multivariate analysis).

|  |  | aOR | 95% CI | p |
| --- | --- | --- | --- | --- |
| Region of birth | France | ref |  | 0.01 |
|  | Maghreb | 1.05 | [0.56-1.96] |  |
|  | Sub-Saharan Africa | 2.66 | [1.46-4.84] |  |
| Health insurance status | SS | ref |  | 0.02 |
|  | CMUc | 1.66 | [0.69-4.16] |  |
|  | AME | 2.10 | [1.11-3.98] |  |
|  | Paid care | 1.65 | [0.74-3.66] |  |
| Combined Occupation category | Other | ref |  | 0.049 |
|  | Not in the labor force or low-skilled occupation* | 1.56 | [1.01-2.42] |  |

\*blue collar or lower white collar

Sensitivity analysis showed that region of birth and being inactive or having a low-skilled occupation were always significantly associated with TNBC (see Supporting Information 2, Tables S2 and S3) whereas being an undocumented migrant was significantly associated with TNBC in only one of the two sensitivity models. All the covariates’ VIF values were below 1.5, suggesting the absence of multicollinearity.

### Frequency of advanced tumors and associated factors

The prevalence of stage T≥3 tumors was also almost 3 times higher in SSA (39.5%, 95% CI = [32.6%-46.9%]) than in FRA (13.4%, 95% CI = [10.1%-17.3 %]) (Figure 2) but they were not significantly different between MAG and FRA (no missing data regarding TNM stage). Prevalence of metastatic stages was not significantly different in the 3 populations: respectively 11.9% in SSA, 8.5% in MAG and 6.3% in FRA (p=0.09).

**Fig 2.**
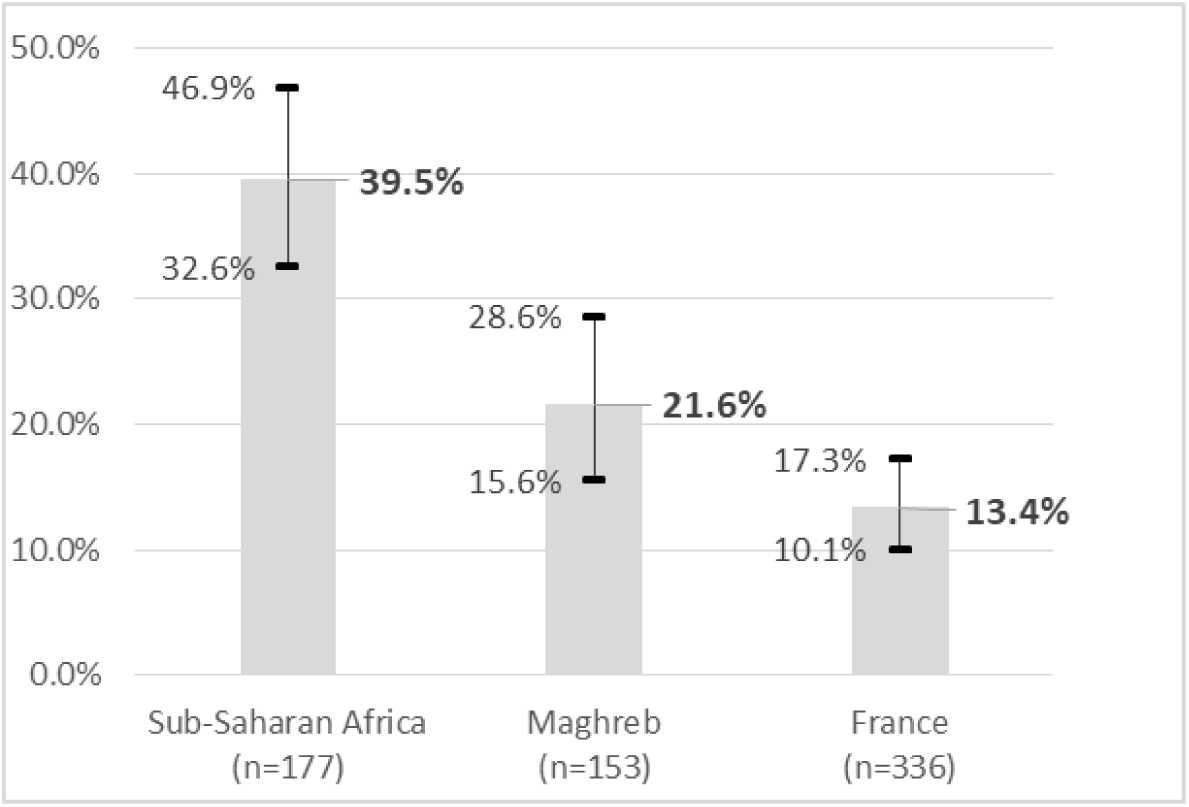
**Frequency of stage T≥3 tumors by country of birth (% and 95% CI).**

Women with a stage T≥3 tumor were significantly younger (on average 5.4 years), with a TNBC, more often SSA, more often undocumented immigrants or foreigners coming from abroad to be treated at Tenon Hospital. (Table 4). There was a correlation between the tumor stage and the absence of markers: 34.7% of the 124 TNBC cases were at stage T≥3, compared to 20.8% of the 486 cases of tumors expressing at least 1 marker (and 12, 7% of the 56 cases with missing marker results, p=0.002).

**Table 4.** Description of stage T≥3 tumors.

|  |  | **T$\geq 3$**<br>n=124 | **T<3**<br>n=486 | **p** |
| --- | --- | --- | --- | --- |
| Age: average (range type) |  | 57.0 (14.9) | 62.4 (12.7) | <0.001 |
| TNBC |  |  |  |  |
|  | Yes | 29.9% | 17.4% |  |
|  | No | 70.1% | 82.6% | 0.001 |
| Region of birth |  |  |  |  |
|  | Sub-Saharan Africa | 47.3% | 20.7% |  |
|  | Maghreb | 22.3% | 23.2% |  |
|  | France | 30.4% | 56.2% | <0.001 |
| Health insurance status |  |  |  |  |
|  | SS | 58.8% | 78.8% |  |
|  | CMUc | 6.8% | 4.2% |  |
|  | AME | 19.6% | 10.8% |  |
|  | Paid care | 14.9% | 6.2% | <0.001 |
| Combined occupation category |  |  |  |  |
|  | Not in the labor force or low-skilled occupation* | 41.2% | 48.3% |  |
|  | Other | 58.8% | 51.7% | 0.077 |
| Combined neighborhood deprivation quintile |  |  |  |  |
|  | <4 | 52.0% | 60.8% |  |
| | $\geq 4$ | 45.3% | 36.5% | |
|  | unknown | 2.7% | 2.7% | 0.151 |

\*blue collar or lower white collar

### Factors affecting tumor stage

In our dataset, differences in tumor stage can be explained by triple negative status but also by a delay in access to health care, which can be due to women’s origin, the quality of their health insurance and/or their poverty. To further investigate the impact of these different factors, we limited our analysis to women residing in France (regularly or not) only; excluding those coming for treatment from abroad (it is likely that they travel to consult in Paris at a more advanced stage, but this is another question). We estimated a logistic regression model that considered age, triple negative status, origin and health insurance status (Table 5). All the covariates’ VIF values were below 1.5, suggesting the absence of multicollinearity. Its estimates showed that triple negative status (in the current model but in neither of the two estimated in the sensitivity analysis: see Supporting Information 2, Tables S4 and S5) and especially the origin of women residing in France were associated with a more advanced tumor stage, but not their insurance status. This last point deserves to be underlined: whether women are insured by usual health insurance (SS), by that for the poor (CMUc) or by that for undocumented immigrants (AME) did not seem to have an influence on the tumor stage at their first consultation.

**Table 5.** Factors associated with stage T≥3.

|  |  | aOR | 95% CI | p |
| --- | --- | --- | --- | --- |
| Age (years) |  | 0.99 | [0.97-1.00] | 0.109 |
| Triple negative | No | Ref |  | 0.034 |
|  | Yes | 1.70 | [1.04-2.79] |  |
| We Region of birth | France | Ref |  | <0.01 |
|  | Maghreb | 1.56 | [0.89-2.72] |  |
|  | Sub-Saharan Africa | 2.85 | [1.58-5.15] |  |
| Health insurance status | SS | Ref |  | 0.70 |
|  | CMUc | 1.19 | [0.50-1.68] |  |
|  | AME | 0.88 | [0.46-1.68] |  |

Women of Sub-Saharan African origin consulted for the first time at the outpatient oncology consultation at a more advanced stage than others, all things being equal regarding the triple negative status of their tumor (and their age). Nearly half of SSA women with TNBC consulted with a tumor at stage T≥3 at the time of diagnosis compared to 27% of French TNBC women (Fig 3). Among non TNBC women, the prevalence of tumors at stage T≥3 was also higher among SSA (37%) than among French women (13%) and this latter difference was statistically significant. Sensitivity analysis led to similar conclusions (see Supporting information, Table S6).

**Fig 3.**
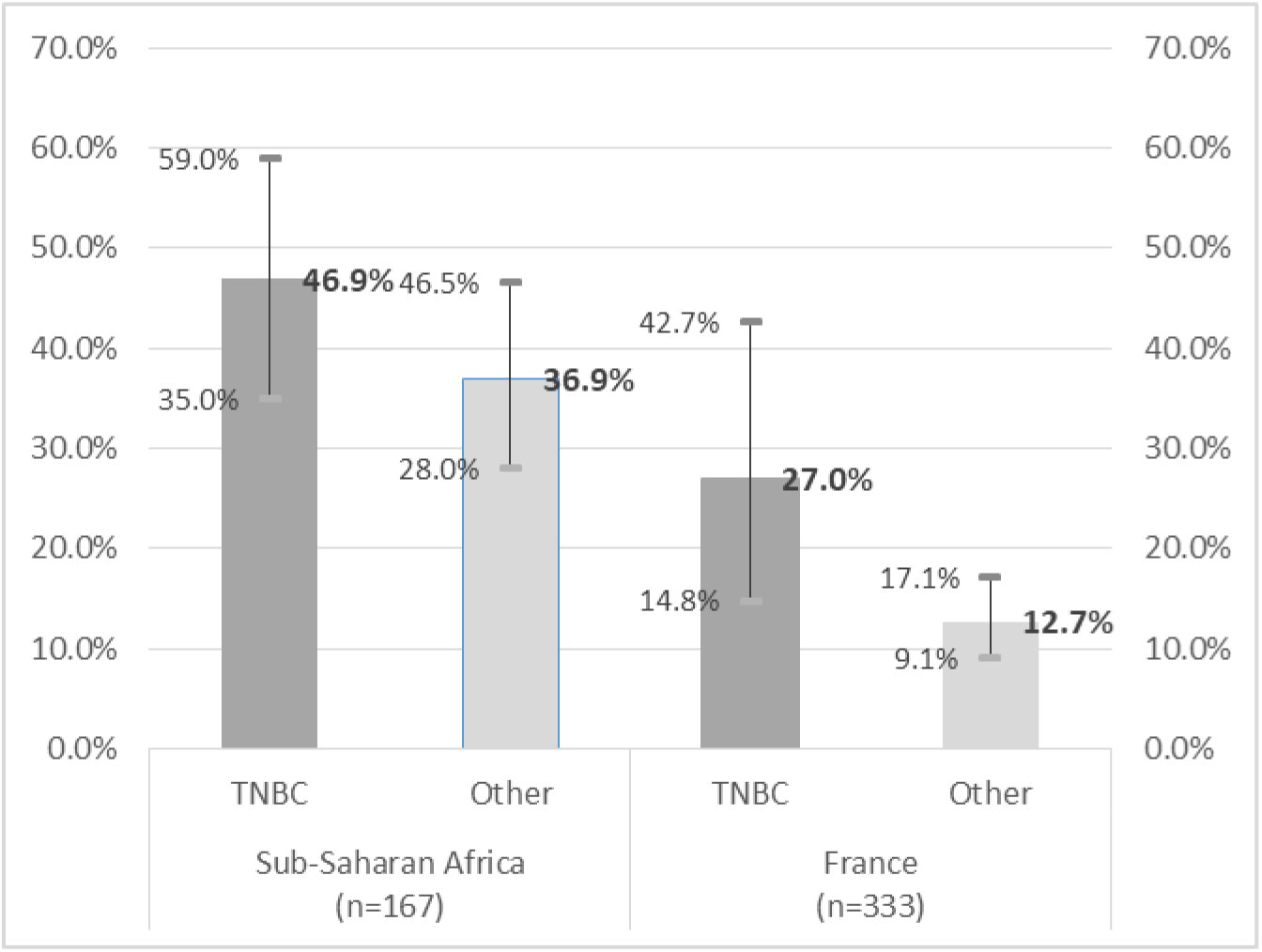
**Comparison of the frequency of stage T≥3 tumors by country of birth according to triple negative status (% and 95% CI).**

## Results of qualitative/ethnographic data

### Factors explaining late access to breast cancer care

This last result can be fruitfully supplemented by certain results from ethnographic research. Indeed, certain cases drew our attention and time. One such example involved a woman in her early thirties with breast cancer with a grim prognosis, whom we visited regularly at home and hospital. She had spent a year consulting an African herbalist living in Paris. He prescribed a medication of stewed leaves to place twice daily on her sore and swollen breast. After deciding that the herbalist’s preparation was not reducing the pain, she finally decided to consult the midwife who had assisted her when she last gave birth. The midwife immediately sent her to a hospital for a mammogram and biopsy. She was subsequently referred to another hospital for chemotherapy, to which she was unresponsive. With metastases and pulmonary problems, she was hospitalized intermittently. Eventually, she returned to Mali, to her husband’s family, to undergo treatment from a ritual specialist. Her mother and her husband’s family agreed that her failure to respond to biomedical treatment was evidence of a “sent” illness, caused by a jealous relative, an etiology often suspected in cases of breast cancer. Her oncologist was supportive of her trip (intended to be a three-week visit) but expressed doubts to the team about the likelihood that she would live to return to France and indeed, she died within two weeks of arriving at the home of the healer.

The pattern evident in the case above, in which the patient waited a year for an herbalist’s treatment to work before turning to biomedical institutions, is similar to that found in other patients’ decision making. For example, Ami came to France from Guinea in response to a job offer from a man she met at work. He offered her a position in a restaurant in Paris, but when she arrived, the job proved to involve sex work. She quit but had no housing so she slept on the street until a social worker in that neighborhood stopped to speak with her and took her to stay with a friend. After settling there, she sought care at a public clinic that accepted undocumented patients to request advice for her painful breast. There, she was referred to a hospital for preliminary examination of her breast condition and then sent to another one for chemotherapy and additional treatment, as is a common process of care seeking. She was told about our team and asked us to accompany her. We found that she was terrified of her diagnosis and convinced that this disease always ends in death. Her immediate problem was that the woman who had agreed to house her required her to move out in case cancer was a contagious disease.

In yet another case, Mariam came to France from Mali to join her younger sister. She had been diagnosed with epilepsy in Mali but was well enough to work as a secretary. She had noticed breast pain and swelling but did not consult anyone for this problem, she explained, because she already had another sickness and did not want to bother her sister with this issue. “Eventually,” months after her arrival in Paris, she told her sister’s general practitioner that she had breast pain. He sent her to a hospital for a mammogram and there she was referred again to another public hospital in the region. Her breast cancer did not respond to chemotherapy and ultimately, she died.

Many patients mentioned the burden of maintaining breast symptoms secret, from the earliest manifestations: a lump, swelling, or pain in the breast. Patients considered it important to hide their concerns from even close family members (including their mother or their husband) to assure that no one in the community would become aware of their symptoms. Several women observed that « *it is very important to keep this sickness (cancer) a secret because ‘Africans’ are capable of ‘doing things’ [faire des choses]*”. By this they meant to engage in evil actions directed at the women concerned, for example ruin their reputation or send a curse to cause sickness. It is very important to emphasize that none of those interviewed mentioned having been informed, in France, of symptoms signifying breast cancer.

### The complexity of cancer care pathways

In general, most patients followed a similar route: first, keeping this anxiety private, then consulting with an herbalist or ritual specialist, or/then getting advice from a social worker or a midwife, and finally referral to a first hospital for mammography and biopsy, the referral to a level 3 hospital certified in oncology. Those for whom mastectomy was recommended but who were undocumented would need to transfer to yet another public hospital, the only one to perform mastectomies on undocumented women. Navigating this complex array of institutions likely constrains some patients from continuing treatment over time. Similarly, the demands of everyday life—collecting a child at day care, trying to maintain a work schedule, domestic responsibilities, traveling long distances from home to the hospital while debilitated from chemotherapy (or in one instance, living in a high-rise apartment on the 14^th^ floor with no functioning elevator): these obligations lead many women to skip appointments or stop treatment altogether.

### What may cause breast cancer?

Among the focus groups held with group leaders and members of immigrant associations, one deserves to be mentioned here. Indeed, in an especially lively discussion, referred to by the 25 participants as a “chat group” (“*une causerie*”), we began by asking a general question: “what causes breast cancer?” Immediately, one woman replied, “money.” Everyone seemed to talk at once, debating whether “money” meant bills or coins. The principal respondent said we (team members) did not seem to appreciate the accommodating capacity of the bra. She proceeded to demonstrate that a cell phone, money (bills and coins) but also a small radio could easily be stored in a woman’s bra. The cell phone and bills were most suspect as causes of breast cancer, given the proximity of the breast to the potentially contaminated objects. In general, the group agreed also that breast cancer, although of uncertain origin, could be a deadly disease sent by a resentful or jealous relative or friend. A co-wife was a particular risk of sending breast cancer to another wife of her husband in polygynous societies.

Given that many West African immigrant women speak only some conversational French, their capacity to comprehend complex explanations of hormone function, tumor growth/stage/grade, metastases, and treatment regimens is limited. In one case, we observed that during over a year of appointments, a particular patient never asked a question of her oncologist. Our data show that many women with breast cancer are misinformed, not informed at all, or disregard medical advice. Not only some patients mention consulting an herbalist or ritual specialist before an appointment with an oncologist as we mentioned above, but others receive packages of herbal preparations from family in West Africa. Also, consultations with healers from Africa living or visiting in Paris may continue throughout the course of hospital interventions. Often clinicians are unaware of this serial or simultaneous reliance on alternative healing remedies.

Accordingly, interpreters had difficulties to translate physicians’ information that they do not understand, and/or consider inappropriate or even dangerous for the patient. For instance, in Bambara, a lingua franca for the region, women we interviewed mentioned that there are numerous words they prefer to use for “cancer” among themselves, such as “*m’bon*” or “*bon*,” (a curse); “*kourou*” or “*kourouni*” (a lump or heaviness); or “*sim di*”. They agreed they prefer not to use the word cancer because - even for well-educated patients - they felt it “*violent*,” and, as many informants told us, it is a word that “*moves towards death*.” They may substitute a religious discourse (for example, “*this is a disease in the hands of God*”) to serve as a less brutal code for delivering “*bad news*”. To say the word cancer, to diagnose cancer and speak of it explicitly to the patient, is a “destiny in itself.”

## Discussion

For the first time in France, a retrospective analysis of hospital medical records such as that presented in this article estimated the frequency of TNBC in women born in Sub-Saharan Africa: it was more than 3 times higher than in women born in France (36.2% versus 11. 0%). The usually reported frequency of 15% of TNBC in mainland France therefore masks very significant differences depending on origins. Should these differences be extrapolated according to race or ethnicity, this gap would be undoubtedly even greater, due to classification biases linked to country of birth (as we mentioned above in the material section). Obviously, some women born in France are of African or Caribbean ancestry, much more frequently than the reverse (i.e. some “White” women may be born in Sub-Saharan Africa). These results are in the same direction as those observed for a long time in the US and the UK, where racial or ethnic health statistics are allowed and conventional. The estimated prevalence among women of African origin in our study is even higher than those estimated in the US among African Americans and among Black women born in West Africa (24%), as well as in women in the French West Indies or in the US black women born in the Caribbean (both around 20%). Our estimated prevalence is close to that estimated in West Africa (where almost all the SSA women in our study were born), which is around 40 to 45%. On the other hand, the prevalence of TNBC among Maghreb immigrant women was not significantly different from that of women born in France and was within the range of estimates published in Maghreb countries [80].

In multivariate analysis, other factors were associated with TNBC: not only African origin (aOR_SSA vs FRA_=2.66) but also belonging to the working class or being out of the labor force (aOR=1.56) and being an undocumented immigrant (aOR=2.10). Since risk estimates in a regression model can be interpreted cumulatively, it means that women meeting these 3 characteristics had a very much higher risk of TNBC. These results are consistent with the mixture of biological, ethno-racial and socioeconomic risk factors of TNBC previously described in the US literature [81]. Many studies have looked at the residential factors of TNBC for decades [82]. Numerous studies actually used the socio-economic status of the neighborhood of residence by default only, as a proxy for the socio-economic status of individuals. More interestingly, others have jointly estimated the role of the ethno-racial category of women and some contextual or environmental factors at the place of residence. We cite a US study which showed the joint influence of race and hazardous environmental exposure at the neighborhood level on the risk of TNBC [83]. Another one, from the same team, showed that both (individual) race and neighborhood concentration of African-American populations were independent risk factors for TNBC, explaining the latter association by a less healthy environment in African-American neighborhoods [84]. On the contrary, others found that higher percentages of African-American residents in the neighborhood were associated with a lower risk of TNBC in African-American women, suggesting that a higher density of the same race/ethnicity may buffer the impacts of disadvantaged neighborhoods [85,86]. In our analyses, we used a neighborhood deprivation index to test a more general psychosocial hypothesis, linked to the social stresses of living in the poorest neighborhoods and its effects on the biology of tumors by altering gene expression [87]. Such a neighborhood effect was not found in our multivariate analysis, but this can be due to low statistical power and/or to a weaker socio-spatial segregation in our population than in those studied in the US.

Our results suggested a later recourse to cancer care, at a more advanced stage, among African women than among women born in France, and this among TNBC patients as well as among others. We have shown that this difference was not associated with patients’ health insurance status, which seems to reflect the egalitarian ideology of the French insurance system. However, this study cannot really judge the equity of the French health care insurance system since it has been carried out in health facilities. Indeed, the enrollment in the CMUc - and even more in the AME - requires complex procedures [88] and it is often done after a medical consultation, with the help of a social worker (in our study one from the hospital cancer department) [89]. Also we know that the rate of non-recourse to the CMUc or the AME could, respectively, reach 30% and even exceed 50% of the eligible populations in France [90]. Ultimately, studying patients’ insurance status in health care settings tells us nothing about the poor, precarious and/or minority populations who do not have access to health care, sometimes because they do not know their rights to health insurance or, once insured, do not know what health care is covered [91]. For instance, we showed in a representative survey conducted in the population of Greater Paris in 2010 that there were strong disparities in delaying or never having undergone breast cancer screening according to origin: being immigrants or born to immigrants were risk factors for delayed and/or no lifetime screening for breast cancer [70]. First, interestingly, this gradient persisted after adjusting for age, health insurance status and socioeconomic status and, second, breast cancer screening is a national, organized, and free of charge program. This is to say that – at least theoretically – there should not exist any significant financial barriers in accessing breast cancer screening or cancer care. So what factors other than financial can therefore come into play, particularly among women of African origin?

The data obtained in the ethnographic study of the experience of breast cancer, supplements and helps to interpret the statistical results. Relatively few studies in France have explored how marginalized populations confront non-communicable illnesses such as cancer [92,93]. In the course of earlier research on immigration, we identified a neglect of chronic, life-threatening illnesses and a lack of data on care taking [94]. Care, in our usage, is a broad concept comprising not only health services but other sectors such as social services, available housing, home caretaking and voluntary association services for structurally vulnerable patients. Immigrant patients who reside in France with serious health problems face not only the biological realities of disease but also the politics of care, which encompass populations [95].

Translation problems dramatically affect patient understandings (even with interpreters, given the absence of relevant vocabulary in patients’ local languages). In Sub-Saharan African countries of origin, awareness and basic biomedical knowledge about cancer are hardly the subject of information campaigns [96]. While some basic knowledge in infectiology and vaccinology has been widely disseminated for decades (as part of expanded vaccination programs, policies to reduce infant mortality, and the fight against tuberculosis and AIDS), and also (may be more recently) about certain chronic diseases such as diabetes or high blood pressure, this is not the case for cancers in general, nor for breast cancer in particular [97]. None of the Sub-Saharan African women we spoke to reported having been informed, in one way or another, about breast cancer since they moved to France (some for decades). “There” as well as “here”, there remains a deep misunderstanding and lack of awareness about breast cancer as a disease [98,99]: what are its causes for modern medicine? where and whom to consult in the health care system for its screening, diagnosis and treatment? [100], what results and prognosis should one expect? [101].

It is therefore not so much the cultural and traditional representations of breast cancer that hinder or delay access to medical care (because it has long been known that medical and traditional representations of illnesses and their treatment can perfectly coexist and we observed it with many African patients that we met [102]), but the almost complete lack of medical information about it dedicated to these minorities groups in France. As a result, one subject remains almost completely unknown to the African women with breast cancer we met: the benefit of early screening (or concern) and to what extent the prognosis is linked to the precocity of the diagnosis. In the absence of medical knowledge, the field of representations seems largely undermined by factors which are obstacles to a request for help and care [103]: fear (including the fear of death), fatalism [104], stigmatization, shame [105], secrecy (including from closest relatives [106]) and self-isolation [107]; not to mention the anticipation of breast mutilation which calls into question the feminine and maternal identities of all the women with breast cancer, but perhaps particularly of African women [108]. The secrecy and hidden conditions of breast cancer has been related in many other studies in women with cancer from ethnic minorities [109], which can prevent patients from sharing their experiences of care and impact their expectations in this matter.

Our study has some limitations. First, even though this is a first in France, the statistical analysis of TNBC by origin is based on a relatively small number of women and on data collected retrospectively from the medical records in an oncology department of a single Parisian hospital. Also, as we mentioned before, our analysis may suffer from classification bias even if such bias cannot fully explain the differences observed. Second, the ethnological results used here come from a broader qualitative research on African women with breast cancer in Greater Paris. By construction, it does not allow for comparison with women of other origins (North African, for example) and the focus was mainly on disadvantaged women. Its results are not generalizable (this was not its goal) to all African patients in Greater Paris. Nevertheless, the women interviewed, like those analyzed in the quantitative study on medical records, have the same social background, residing in working-class, often underserved, neighborhoods in eastern Paris. Third, our mixed methods survey is sequential in a way that may seem frustrating. Indeed, one would have dreamed of supplementing it with a subsequent quantitative survey examining many more social and cultural variables than those available in medical records. On the one hand, many ethnological results constitute hypotheses that could be statistically confirmed; on the other hand, women’s socioeconomic characteristics could be much more detailed (socioeconomic status but also length of stay in France, fluency in French, social isolation, family and social support, etc. [110,111]). Finally, ideally, such a future study—necessarily prospective—should be multicentered. This means that more ambitious research remains to be undertaken in the French context.

## Conclusion

Described as a public health emergency in the United States [112], breast cancer in Black women is not the subject of any specific public health recommendations in France. The absence of ethno-racial data in French cancer registries and medical records prevents any large-scale study both on ethnic epidemiology of TNBC, and on discrimination and its role in breast cancer disparities, a subject recently deemed a priority by the Lancet Breast Cancer Commission [113]. Breast cancers’ clinical and epidemiological specificities in Black women, in particular those with TNBC call for specific therapeutic strategies and, in the case of immigrant women from Sub-Saharan Africa in France, culturally adapted information and support. Recently, some authors have even suggested that the epidemiological characteristics of breast cancer in Black women should lead to advancing the target age for organized screening by 8 years for them compared to White women of the majority population [114]. In a country like France, and beyond in all European countries, so much attached to equal access to care and to health equity through public health systems or public health insurance as one of the pillars of their welfare states, public health surveillance, information, prevention, screening and care of breast cancers altogether need to be specifically adapted according to women’s histories, lived experiences, and ethno-racial belonging.

The epidemiological and ethnographic data presented here convey the ways in which regulations and conventions regarding public discourse on race and ethnicity in France and other European nations constrain clear social scientific evaluation of illness diagnosis and treatment. Racism is a contested and emotionally fraught subject in France, as in so many other societies. In spite of the emphasis by politicians, public intellectuals, and the French public at large on equality (illustrated by race and ethnic blindness), our data unpacks what must be regarded as difficult realities regarding ethno-racial identities. The goal of this article is to advance public knowledge and discourse, to elucidate the decisions made by people facing life-threatening disease, and to show that their agency is informed by multiple factors—social relations in their home societies, population genetics, economic constraints, and the impact of state policies in France.

## Data Availability

Minimal dataset that can be publicly accessed in Zenodo depository.

https://doi.org/10.5281/zenodo.17226211

## Acknowledgments

The authors gratefully acknowledge Professor Sarah Gehlert at Washington University in Saint Louis for her expertise on ethnoracial disparities in breast cancer in the United States. They also thank Professor Laurent Zelek at Avicennes Hospital (APHP, Bobigny) for his long-standing involvement in the ethnographic study reported in this article.

## Supporting information 1. McGill Illness Narrative Interview (MINI)

Groleau D, Young A, Kirmayer LJ. The McGill Illness Narrative Interview (MINI): an interview schedule to elicit meanings and modes of reasoning related to illness experience. *Transcult Psychiatry*. 2006 Dec;43(4):671-91. doi: 10.1177/1363461506070796.

**Section 1. INITIAL ILLNESS NARRATIVE**

1. When did you experience your health problem or difficulties (HP) for the first time? [*Substitute respondent’s terms for ‘HP’ in this and subsequent questions.*] [*Let the narrative go on as long as possible, with only simple prompting by asking*, ‘What happened then? And then?’]
2. We would like to know more about your experience. Could you tell us when you realized you had this (HP)?
3. Can you tell us what happened when you had your (HP)?
4. Did something else happen? [*Repeat as needed to draw out contiguous experiences and events.*]
5. If you went to see a helper or healer of any kind, tell us about your visit and what happened afterwards.
6. If you went to see a doctor, tell us about your visit to the doctor/hospitalization and about what happened afterwards.

6.1 Did you have any tests or treatments for your (HP)? [The relevance of this question depends on the type of health problem.]

**Section 2. PROTOTYPE NARRATIVE**

7. In the past, have you ever had a health problem that you consider similar to your current (HP)? [If answer to #7 is Yes, then ask Q.8]
8. In what way is that past health problem similar to or different from your current (HP)?
9. Did a person in your family ever experience a health problem similar to yours? [If answer to #9 is Yes, then ask Q.10]
10. In what ways do you consider your (HP) to be similar to or different from this other person’s health problem?
11. Did a person in your social environment (friends or work) experience a health problem similar to yours? [If answer to #11 is Yes, then ask Q.12]
12. In what ways do you consider your (HP) to be similar to or different from this other person’s health problem?
13. Have you ever seen, read or heard on television, radio, in a magazine, a book or on the Internet of a person who had the same health problem as you? [If answer to #13 is Yes, then ask Q.14]
14. In what ways is that person’s problem similar to or different from yours?

**Section 3. EXPLANATORY MODEL NARRATIVE**

15. Do you have another term or expression that describes your (HP)?
16. According to you, what caused your (HP)? [List primary cause(s).]

16.1 Are there any other causes that you think played a role? [List secondary causes.]

17. Why did your (HP) start when it did?
18. What happened inside your body that could explain your (HP)?
19. Is there something happening in your family, at work or in your social life that could explain your health problem? [If answer to #19 is Yes, then ask Q.20]
20. Can you tell me how that explains your health problem?
21. Have you considered that you might have *[INTRODUCE POPULAR SYMPTOM OR ILLNESS LABEL]*?
22. What does *[POPULAR LABEL]* mean to you?
23. What usually happens to people who have *[POPULAR LABEL]?*
24. What is the best treatment for people who have *[POPULAR LABEL]?*
25. How do other people react to someone who has *[POPULAR LABEL]?*
26. Who do you know who has had *[POPULAR LABEL]*?
27. In what ways is your (HP) similar to or different from that person’s health problem?
28. Is your (HP) somehow linked or related to specific events that occurred in your life?
29. Can you tell me more about those events and how they are linked to your (HP)?

**Section 4. SERVICES AND RESPONSE TO TREATMENT**

30. During your visit to the doctor (healer) for your HP, what did your doctor (healer) tell you that your problem was?
31. Did your doctor (healer) give you any treatment, medicine or recommendations to follow? [List all]
32. How are you dealing with each of these recommendations? [*Repeat Q. 33 to Q. 36 as needed for every recommendation, medicine and treatment listed.*]
33. Are you able to follow that treatment (or recommendation or medicine)?
34. What made that treatment work well?
35. What made that treatment difficult to follow or work poorly?
36. What treatments did you expect to receive for your (HP) that you did not receive?
37. What other therapy, treatment, help or care have you sought out?
38. What other therapy, treatment, help or care would you like to receive?

**Section 5. IMPACT ON LIFE**

39. How has your (HP) changed the way you live?
40. How has your (HP) changed the way you feel or think about yourself?
41. How has your (HP) changed the way you look at life in general?
42. How has your (HP) changed the way that others look at you?
43. What has helped you through this period in your life?
44. How have your family or friends helped you through this difficult period of your life?
45. How has your spiritual life, faith or religious practice helped you go through this difficult period of your life?
46. Is there anything else you would like to add?

**Supporting information 2. Sensitivity analysis for missing tumor markers data.**

Due to missing data for one or more tumor markers, triple-negative status was unknown for 56 women: 10 of 177 SSA women (5.6%), 14 of 153 MAG (9.2%), and 3 of 336 FRA (0.9%), p<0.001.

A sensitivity analysis was performed, assuming all missing values were positive or negative, respectively.

---

**Table S1.** Sensitivity analysis for the prevalence of TNBC.

|  | Sub-Saharan Africa | Maghreb | France |
| --- | --- | --- | --- |
| **Results reported\*** | n=167 | n=142 | n=333 |
| TNBC | 38.3% | 16.5% | 12.2% |
| 95%CI | [31.2%-45.8%] | [11.1%-23.4%] | [8.9%-16.2%] |
| **Sensitivity analysis** | n=177 | n=153 | n=336 |
| *if all missing values for triple negative status = "yes"* |  |  |  |
| TNBC | 41.8% | 24.2% | 20.5% |
| 95%CI | [34.7%-49.2%] | [17.9%-31.4%] | [16.5%-25.1%] |
| *if all missing values for triple negative status = "no"* |  |  |  |
| TNBC | 36.2% | 15.0% | 11.0% |
| 95%CI | [29.4%-43.4%] | [10.0%-21.3%] | [8.0%-14.7%] |

\*referring to Figure 1 of the main article

**Table S2.** Factors associated with TNBC (multivariate analysis), if all missing values for triple negative status = “yes”.

|  |  | aOR | 95% CI | p |
| --- | --- | --- | --- | --- |
| Region of birth | France | ref |  | 0.036 |
|  | Maghreb | 0.94 | [0.57-1.55] |  |
|  | Sub-Saharan Africa | 1.74 | [1.03-2.94] |  |
| Health insurance status | SS | ref |  | 0.07 |
|  | CMUc | 1.60 | [0.73-3.65] |  |
|  | AME | 2.16 | [1.20-3.86] |  |
|  | Paid care | 1.39 | [0.70-2.77] |  |
| Combined Occupation category | Other | ref |  | 0.005 |
|  | Not in the labor force or low-skilled occupation* | 1.77 | [1.03-2.94] |  |

\*blue collar or lower white collar

**Table S3.** Factors associated with TNBC (multivariate analysis), if all missing values for triple negative status = “no”.

|  |  | aOR | 95% CI | p |
| --- | --- | --- | --- | --- |
| Region of birth | France | ref |  | <0.001 |
|  | Maghreb | 1.15 | [0.63-2.11] |  |
|  | Sub-Saharan Africa | 3.19 | [1.77-5.73] |  |
| Health insurance status | SS | ref |  | 0.38 |
|  | CMUc | 1.45 | [0.60-3.50] |  |
|  | AME | 1.73 | [0.93-3.22] |  |
|  | Paid care | 1.36 | [0.65-2.85] |  |
| Combined Occupation category | Other | ref |  | 0.03 |
|  | Not in the labor force or low-skilled occupation* | 1.60 | [1.04-2.46] |  |

\*blue collar or lower white collar

**Table S4.** Factors associated with stage T≥3 (multivariate analysis), if all missing values for triple negative status = “yes”.

|  |  | aOR | 95% CI | p |
| --- | --- | --- | --- | --- |
| Age (years) |  | 0.99 | [0.97-1.0] | 0.07 |
| Triple negative | No | Ref |  | 0.98 |
|  | Yes | 1.00 | [0.66-1.55] |  |
| Region of birth | France | Ref |  | <0.001 |
|  | Maghreb | 1.56 | [0.91-2.65] |  |
|  | Sub-Saharan Africa | 3.25 | [1.86-5.67] |  |
| Health insurance status | SS | Ref |  | 0.68 |
|  | CMUc | 1.10 | [0.47-2.60] |  |
|  | AME | 1.00 | [0.54-1.84] |  |

**Table S5.** Factors associated with stage T≥3 (multivariate analysis), if all missing values for triple negative status = “no”.

|  |  | aOR | 95% CI | p |
| --- | --- | --- | --- | --- |
| Age (years) |  | 0.99 | [0.97-1.00] | 0.09 |
| Triple negative | No | Ref |  | 0.29 |
|  | Yes | 1.28 | [0.81-2.03] |  |
| Region of birth | France | Ref |  | <0.001 |
|  | Maghreb | 1.75 | [1.00-3.03] |  |
|  | Sub-Saharan Africa | 3.17 | [1.78-5.65] |  |
| Health insurance status | SS | Ref |  | 0.98 |
|  | CMUc | 1.04 | [0.44-2.44] |  |
|  | AME | 0.95 | [0.51-1.76] |  |

**Table S6.** Sensitivity analysis for the frequency of stage T≥3 tumors by country of birth according to triple negative status.

|  | Sub-Saharan Africa |  | France |  |
| --- | --- | --- | --- | --- |
|  | TNBC | Other | TNBC | Other |
| **Results reported** | n=167 |  | n=333 |  |
| T>=3 | 46.9% | 36.9% | 27.0% | 12.7% |
| 95%CI | [35.0%-59.0%] | [28.0%-46.5%] | [14.8%-42.7%] | [9.1%-17.1%] |
| **Sensitivity analysis** | n=177 |  | n=336 |  |
| *if all missing values for triple negative status = "yes"* |  |  |  |  |
| T>=3 | 43.2% | 36.9% | 15.9% | 12.7% |
| 95%CI | [32.4%-54.6%] | [28.0%-46.5%] | [8.8%-25.9%] | [9.1%-17.1%] |
| *if all missing values for triple negative status = "no"* |  |  |  |  |
| T>=3 | 46.9% | 35.4% | 27.0% | 11.7% |
| 95%CI | [35.0%-59.0%] | [27.0%-44.5%] | [14.8%-42.7%] | [8.4%-15.7%] |

*\*referring to Figure 3 of the main article*

## References

1 Boyle P. Triple negative breast cancer: epidemiological considerations and recommendations. Ann Oncol. August 2012;23(S6):7–12.

2 Cortet M, Bertaut A, Molinié F et al. Trends in molecular subtypes of breast cancer: description of incidence rates between 2007 and 2012 from three French registries. Cancer BMC. 2018;18:161.

3 Hassaine Y, Jacquet E, Seigneurin A et al. Evolution of the incidence of breast cancer in young women in a French registry from 1990 to 2018: towards a change in screening strategy? Breast Cancer Res. 2022;24(1):87.

4 Richardson LC, Henley SJ, Miller JW, Massetti G, Thomas CC. Patterns and Trends in Age-Specific Black-White Differences in Breast Cancer Incidence and Mortality - United States, 1999-2014. MMWR Morb Mortal Wkly Rep. 2016 Oct 14;65(40):1093–98.

5 Edit K, Hicks D, Ambrosone CB. Breast cancer in African-American women: differences in tumor biology compared to European-American women. Cancer Res. September 1, 2006;66(17):8327–30.

6 Gordon NH, Crowe JP, Brumberg DJ, Berger NA. Socioeconomic factors and race in breast cancer recurrence and survival. Am J Epidemiol. March 15, 1992;135(6): 609–18.

7 Newman LA, Kaljee LM. Health disparities and triple negative breast cancer among African American women: a review. JAMA Surg. 2017;152(5):485–93.

8 Prakash O, Hossain, Danos, Lassak A, Scribner R, Miele L. Racial disparities in triple-negative breast cancer: an examination of the role of biological and non-biological factors. Public Health Front. Dec 2020;8:576964.

9 John EM, Miron A, Gong G et al. Prevalence of pathogenic BRCA1 mutation carriers in 5 racial/ethnic groups in the United States. JAMA. 2007;298:2869–76.

10 Malone KE, Daling JR, Doody DR, et al. Prevalence and predictors of BRCA1 and BRCA2 mutations in a population-based study of breast cancer in white and black American women ages 35 to 64 years. Cancer Res. 2006 Aug 15;66(16):8297–308.

11 Stark A, Kleer CG, Martin I et al. African ancestry and higher prevalence of triple negative breast cancer: results of an international study. Cancer. 2010;116:4926–32.

12 Quan L, Gong Z, Yao S et al. Adaptive immune response cytokine and cytokine receptor genes are differentially associated with breast cancer risk in American women of African and European ancestry. Int J Cancer. 2014;134:1408–21.

13 Howard FM, Olopade OI. Epidemiology of Triple-Negative Breast Cancer. Cancer J. 2021;27(1):8–16.

14 Sung H, DeSantis CE, Fedewa SA, Kantelhardt EJ, Jemal A. Breast cancer subtypes among Eastern-African-born black women and other black women in the United States. Cancer. 2019 Oct 1;125(19):3401–11.

15 Newman LA, Griffith KA, Jatoi I, Simon MS, Crowe JP, Colditz GA. Meta-analysis of survival in African American and White American breast cancer patients: ethnicity compared with socioeconomic status. J Clin Oncol. March 20, 2006;24(9):1342–9.

16 Hudson D, Gehlert S. Considering the role of social determinants of health in disparities between black and white breast cancer. In: Bangs R, Davis LE, eds. Racial and Social Issues: Restructuring Inequalities. New York: Springer Press, 2014: 227-46.

17 Carey LA, Peru CM, Livasy CA et al. Race, breast cancer subtypes, and survival in the Carolina Breast Cancer Study. JAMA. June 7, 2006;295(21):2492–502.

18 Dietze EC, Sistrunk C, Miranda-Carboni G, O’Regan R, Seewaldt VL. Triple-negative breast cancer in African-American women: disparities versus biology. Nat Rev Cancer. 2015 April;15 (4):248–54.

19 Siddharth S, Parida S, Muniraj N et al. Concomitant activation of GLI1 and Notch1 contributes to the racial disparity in human triple negative breast cancer progression. Elife. December 10, 2021;10:e70729.

20 Linnenbringer E, Gehlert S, Geronimus A. Black-white disparities in breast cancer subtype: the intersection of socially structured stress and gene expression. AIMS Public Health. 2017;4(5):526–56.

21 Jack RH, Davies EA, Renshaw C, Tutt A, Grocock MJ, Coupland VH, Møller H. Differences in breast cancer hormone receptor status across ethnic groups: a London population. Eur J Cancer. February 2013;49(3):696–702.

22 Sandhu GS, Erqou S, Patterson H, Mathew A. Prevalence of triple negative breast cancer in India: systematic review and meta-analysis. J Glob Oncol. June 29, 2016;2(6):412–21.

23 Thakur KK, Bordoloi D, Kunnumakkara AB. Alarming burden of triple negative breast cancer in India. Clin Breast Cancer. June 2018;18(3):e393–9.

24 Copson E, Maishman T, Gerty S et al. Ethnic origin and outcome of young breast cancer patients in the United Kingdom: the POSH study. Br J Cancer. January 7,2014;110(1):230–41.

25 Gathani T, Reeves G, Broggio J, Barnes I. Ethnicity and tumor characteristics of invasive breast cancer in over 116,500 women in England. Br J Cancer. August 2021;125(4):611–17.

26 Fry A, White B, Nagarwalla D, Shelton J, Jack RH. Relationship between ethnicity and stage at diagnosis in England: a national analysis of six cancer sites. BMJ Open. January 26, 2023;13(1):e062079.

27 Forbes LJ, Atkins L, Thurnham A, Layburn J, Haste F, Ramirez AJ. Breast cancer awareness and barriers to symptomatic presentation among women from different ethnic groups in East London. Br J Cancer. November 8,2011;105(10):1474–9.

28 Waller J, Robb K, Stubbings S et al. Awareness of cancer symptoms and anticipated help-seeking among ethnic minority groups in England. Br J Cancer. December 3, 2009;101(S2): S24–30.

29 Vrinten, Wardle J, Marlow LA. Fear of cancer and fatalism among ethnic minority women in the United Kingdom. Br J Cancer. 2016;114:597–604.

30 Niksic M, Rachet B, Warburton FG, Forbes LJ. Ethnic differences in awareness of cancer symptoms and barriers to seeking medical help in England. Br J Cancer. 2016 Jun 28;115(1):136–44.

31 Adeloye D, Sowunmi OY, Jacobs W et al. Estimation of breast cancer incidence in Africa: a systematic review and meta-analysis. J Glob Health. June 2018;8(1):010419.

32 Ziegenhorn HV, Frie KG, Ekanem IO et al. Breast cancer pathology services in sub-Saharan Africa: a survey within population-based cancer registries. BMC Health Serv Res. 2020 Oct 2;20(1):912.

33 Onyia AF, Nana TA, Adewale EA et al. Breast cancer phenotypes in Africa: scoping review and meta-analysis. JCO Globe Oncol. September 2023;9:e2300135.

34 Hercules SM, Alnajar M, Chen C et al. Prevalence of triple negative breast cancer in Africa: a systematic review and meta-analysis. BMJ Open. May 27, 2022;12(5):e055735.

35 Jiagge E, Oppong JK, Bensenhaver J, et al. Breast Cancer and African Ancestry: Lessons Learned at the 10-Year Anniversary of the Ghana-Michigan Research Partnership and International Breast Registry. J Glob Oncol. 2016 Jul 27;2(5):302–310.

36 Ly M, Antoine M, Dembélé AK, et al. High incidence of triple negative tumors in sub-Saharan Africa: a prospective study of the characteristics and risk factors of breast cancer in Malian women seen in a university hospital in Bamako. Oncology. 2012;83(5):257–63.

37 Fitzpatrick MB, Rendi MH, Kiviat NB, et al. Pathology of Senegalese breast cancers. Pan Afr Med J. October 2, 2019;34:67.

38 Aka E, Horo A, Koffi A, Fanny M, Didi-Kouko C, Nda G, Abouna A, Kone M. [Breast cancer management in Abidjan: A single-center experience]. Gynecol Obstet Fertil Senol. September 2021;49(9):684–90.

39 Adani-Ifè A, Amégbor K, Doh K, Darré T. Breast cancer in Togolese women: immunohistochemistry subtypes. BMC Women Health. November 23, 2020;20(1):261.

40 Parenté A, Gnangnon F, Ivanga M, Sacca H, Laleye A, Darré T, et al. Le cancer du sein en Afrique subsaharienne, un nouveau défi sanitaire. Rev Épidémiol Santé Publique. 2023;71(S3): 101893.

41 Traoré B, Koulibaly M, Diallo A, Bah M. Molecular profile of breast cancers in Guinean oncological environment. Pan Afr Med J. May 14, 2019;33:22.

42 Biancodella M, Ouédraogo NLM, Zongo N, et al. Breast cancer in West Africa: molecular analysis of BRCA genes in early-onset breats cancer patients in Burkina Faso. Hum Genomics. 2021 Oct 30;15(1):65.

43 Fackenthal JD, Zhang J, Zhang B, Zheng Y, Hagos F, Burrill DR, Niu Q, Huo D, Sveen WE, Ogundiran T, Adebamowo C, Odetunde A, Falusi AG, Olopade OI. High prevalence of BRCA1 and BRCA2 mutations in unselected Nigerian breast cancer patients. Int J Cancer. 2012 Sep 1;131(5):1114–23.

44 Ugiagbe EE, Owolabi DO. Molecular Subtypes of Breast Cancer in a Tertiary Centre in Edo State: South-South Nigeria. West Afr J Med. 2024 Jul 30;41(7):767–774.

45 Igibah CO, Asogun D, Omonfuegbe OM, et al. Distribution of Immunohistochemical Subtypes of Breast Cancer in Nigeria: A Systematic review. Cureus. 2025 Sep 7;17(9):e91806.

46 Osagie K. The return of the myth of race? New Scientist. 2009;203(2715):22–3.

47 Qiang V, Tope P, Roberge C, Britnei J, Elmi Y, Leggett S. Race-based data alone is not enough: a call to action. Lancet. 2024;403(10423):228–31.

48 Public Health England. Local action on health inequalities: understanding and reducing ethnic inequalities in health. https://www.gov.uk/government/publications/health-inequalities-reducing-ethnic-inequalities (2018).

49 Chauvin P. Interroger la situation sociale des individus. *In*: Vuillermoz C et al, eds. L’épidémiologie sociale: concepts, méthodes et exemples d’utilisation. Rennes: Presses de l’EHESP, 2024: 57–68.

50 Lévi-Strauss C. Race et Histoire. Paris: Folio, 1989 (édition originale 1952): 10.

51 Jacquard A. In praise of difference: genetics and human affairs. New York: Columbia University Press, 1984.

52 Lainé A. L’anthropologie biologique et l’Afrique au XXe siècle. In: Deslauriers C, Juhé-Beaulaton D, eds. Afrique, terre d’histoire. Sur les traces de Jean-Pierre Chrétien. Paris: Karthala, 2007: 131-58.

53 Héran V. France/Etats-Unis: deux visions de la statistique des origines et des minorités ethniques. Santé Société et Solidarité. 2005;(1): 167–89.

54 Fauquet G. Note sur la population de la Martinique (éléments ethniques et catégories sociales). Bulletins et Mémoires de la Société d’anthropologie de Paris. 1912;VI-3(2):154-61.

55 Fassin E, Fassin D. De la question sociale à la question raciale: Représenter la société française. Paris: La Découverte, 2006.

56 Simon P. The Choice of Ignorance: The Debate on Ethnic and Racial Statistics in France. *In*: Simon P, Piché V, Gagnon A, eds. Social Statistics and Ethnic Diversity. IMISCOE Research Series. Springer, Cham, 2015: 65–87.

57 Héran F. Cessons d’opposer les principes républicains à la statistique ethnique. Le Monde, 24/06/2020.

58 Azria E, Sauvegrain P, Blanc J, et al. Racisme systémique et inégalités de santé, une urgence sanitaire et sociétale révélée par la pandémie COVID-19 [Systemic racism and health inequalities, a sanitary emergency revealed by the COVID-19 pandemic]. Gynecol Obstet Fertil Senol. 2020 Dec;48(12):847–49.

59 Melchior M, Desgrées du Loû A, Gosselin A, et al. À quand une prise en compte des disparités ethnoraciales vis-à-vis de l’infection à COVID-19 en France ? [Advocacy for the measurement and reduction of COVID-19 disparities according to migratory status and ethnic category in France]. Rev Epidemiol Sante Publique. 2021 Apr;69(2):96–8.

60 Lê J, Simon P, Coulmont B. La diversité des origines et la mixité des unions progressent au fil des générations. Insee Première n°1910, juillet 2022.

61 Beauchemin C, Hamel C, Lesné M, Simon P. Les discriminations: une question de minorités visibles. Population & Sociétés. 2010;Avril(466):1-4.

62 McAvay H, Safi M. Class versus race? Multidimensional inequality and intersectional identities in France. Ethn Racial Stud. 2023;46(15):3167–98.

63 Combien y a-t-il de Noirs en FranceAlternatives Economiques, février 2008, Hors-série n° 076.

64 Sabeg Y, Méhaignerie L. Les oubliés de l’égalité des chances. Participation, pluralité, assimilation…ou repli *?* Paris: Institut Montaigne, 2004.

65 Barou J. Les immigrations africaines en France au tournant du siècle. Hommes & Migrations. 2002;1239:6–18.

66 Timera M, Garnier, J. (2010). Les Africains en France Vieillissement et transformation d’une migration. Hommes & Migrations. 2010; 1286-1287(4): 24–35.

67 Bidoux PE. Les descendants d’immigrés se sentent au moins autant discriminés que les immigrés. Insee Ile-de-France. Octobre 2012, n°395.

68 Zander U. La hiérarchie « socio-raciale » en Martinique. Entre persistances postcoloniales et évolution vers un désir de vivre ensemble. Revue Asylon(s), mai 2013, n°11.

69 Beaubrun-Renard MM, Véronique-Baudin J, Macni J, et al. Overall Survival of triple negative breast cancer in French West Indian women. PLoS One. August 24, 2022;17(8):e0271966.

70 Deloumeaux J, Gaumond S, Bhakkan B et al. Incidence, mortality and receptor status of breast cancer in Afro-Caribbean women: Data from the Guadeloupe cancer registry. Cancer Epidemiol. April 2017;47:42–7.

71 Poiseuil M, Coureau G, Payet C, et al. Deprivation and mass screening: Survival of women diagnosed with breast cancer in France from 2008 to 2010. Cancer Epidemiol. June 2019;60:149–55.

72 Bryc K, Durand EY, Macpherson JM, Reich D, Mountain JL. The genetic ancestry of African Americans, Latinos, and European Americans across the United States. Am J Hum Genet. 2015 Jan 8;96(1):37–53.

73 Beauchemin C, Hamel V, Simon, Cris P, eds. Trajectoires et origines. Enquête sur la diversité des populations en France. Paris: Ined Éditions, 2016.

74 Desgrées du Loû, Lert F, eds. Parcours de vie et santé des Africains immigrées en France. Paris: La Découverte, 2017.

75 Rondet C, Lapostolle A, Soler M, Grillo F, Parizot I, Chauvin P. Are immigrants and nationals born to immigrants at higher risk for delayed or no lifetime breast and cervical cancer screening? The results from a population-based survey in Paris metropolitan area in 2010. *PLoS One*. 2014 Jan 22;9(1):e87046.

76 Rey G, Jougla E, Fouillet A, Hémon D Ecological association between an index of deprivation and mortality in France over the period 1997 - 2001: variations according to spatial scale, degree of urbanity, age, sex and the cause of death. BMC Public Health. January 22, 2009;9:33.

77 Sargent C. The Navigation of Public Hospitals by West African Immigrants in Paris, France. In: Olsen WC, Sargent C, eds. The Work of Hospitals. Global Medicine in Local Cultures. New Brunswick: Rutgers University Press, 2022, pp. 165-81.

78 Sargent C, Zelek L, Festa A. Precarity, Chronic Illness, and Borders of Care Confronting Immigrants in Paris, France. In: El-Shaarawi N, Larchanché S, eds. Migration and Health: Challenging the Borders of Belonging, Care, and Policy. New York, Oxford: Berghahn Books, 2022, pp. 124-38.

79 Groleau D, Young A, Kirmayer LJ. The McGill Illness Narrative Interview (MINI): an interview schedule to elicit meanings and modes of reasoning related to illness experience. Transcult Psychiatry. 2006 Dec;43(4):671–91.

80 Corbex M, Bouzbid S, Boffetta P. Features of breast cancer in developing countries, examples from North-Africa. Eur J Cancer. 2014 Jul;50(10):1808–118.

81 Llanos AA, Chandwani S, Bandera EV, Hirshfield KM, Lin Y, Ambrosone CB, et al. Association between sociodemographic and clinicopathological factors and breast cancer subtypes in a population-based-study. Cancer Causes Control. 2015;26:1737–50.

82 Krieger N. Social class and the black/white crossover in the age-specific incidence of breast cancer: a study linking census-derived data to population-based registry records. Am J Epidemiol. 1990 May;131(5):804–14.

83 Siegel SD, Brooks MM, Berman JD, et al. Neighborhood factors and triple negative breast cancer: The role of cumulative exposure to area-level risk factors. Cancer Med. 2023 May;12(10):11760–72.

84 Siegel SD, Brooks MM, Lynch SM, Sims-Mourtada J, Schug ZT, Curriero FC. Racial disparities in triple negative breast cancer: toward a causal architecture approach. Breast Cancer Res. 2022 Jun 1;24(1):37.

85 Linnenbringer E, Geronimus AT, Davis KL, Bound J, Ellis L, Gomez SL. Associations between breast cancer subtype and neighborhood socioeconomic and racial composition among Black and White women. Breast Cancer Res Treat. 2020 Apr;180(2):437–47.

86 Qin B, Babel RA, Plascak JJ, Lin Y, Stroup AM, Goldman N, Ambrosone CB, Demissie K, Hong CC, Bandera EV, Llanos AAM. Neighborhood Social Environmental Factors and Breast Cancer Subtypes among Black Women. Cancer Epidemiol Biomarkers Prev. 2021 Feb;30(2):344–50.

87 Antonova L, Aronson K, Mueller CR. Stress and breast cancer: from epidemiology to molecular biology. Breast Cancer Res. 2011 Apr 21;13(2):208

88 Sargent C, Kotobi L. Austerity and its implications for immigrant health in France. Soc Sci Med. 2017 Aug;187:259–67.

89 Pian A. De l’accès aux soins aux « trajectoires du mourir ». Les étrangers atteints de cancer face aux contraintes administratives. Rev Eur Migr Int (REMI). 2012 Oct;28(2): 101–27.

90 Dourgnon P, Jusot F, Marsaudon A, Sarhiri J, Wittwer J. Just a question of time? Explaining non-take-up of a public health insurance program designed for undocumented immigrants living in France. Health Econ Policy Law. 2023 Jan;18(1):32–48.

91 Silber JH, Rosenbaum PR, Ross RN, et al. Disparities in Breast Cancer Survival by Socioeconomic Status Despite Medicare and Medicaid Insurance. Milbank Q. 2018 Dec;96(4):706–54.

92 Pian A. De l’accès aux soins aux « trajectoires du mourir ». Les étrangers atteints de cancer face aux contraintes administratives. Rev Eur Migr Int. 2012; 28(2): 101–27.

93 Sarradon-Eck A. Le cancer comme inscription d’une rupture biographique dans le corps. In: Cousson-Gélie F, Langlois E, Barrault M, eds. Faire face au cancer. Image du corps, image de soi. Paris: Tikinagan, 2009, pp. 285-311.

94 Kotobi L, Sargent C. Precarity and cancer among low-income populations in France: intractable inequalities. In: Benett LR, Manderson L, Spagnoletti B, eds. Cancer and the Politics of Care: Inequalities and Interventions in Global Perspective. London, UCL Press, 2023, pp. 232–253.

95 Larchanché S. Cultural Anxieties. Managing Migrant Suffering in France. New Brunswick: Rutgers University Press, 2020, pp. 187-188.

96 Schantz C, Coulibaly A, Traoré A, Traoré BA, Faye K, Robin J, et al. Access to oncology care in Mali: a qualitative study on breast cancer. BMC Cancer. 2024;24:81.

97 Livingston J. Improvising Medicine. An African Oncoloy Ward in an Emerging Cancer Epidemic. Durham: Duke University Press, 2012, pp. 51-5.

98 Ly M, Diop S, Sacko M, Baby M, Diop CT, Diallo DA. Cancer du sein: facteurs influençant l’itinéraire thérapeutique des usagers d’un service d’oncologie médicale à Bamako (Mali) [Breast cancer: factors influencing the therapeutic itinerary of patients in a medical oncology unit in Bamako (Mali)]. Bull Cancer. 2002;89:323–26.

99 Sulu SMM, Mukuku O, Sulu AMS, Massamba BL, Mashinda DK, Tshimpi AW. Knowledge regarding breast cancer among Congolese women in Kinshasa, Democratic Republic of the Congo. Cancer Rep (Hoboken). 2023 Mar;6(3):e1758.

100 Berdzuli N. Breast cancer: from awareness to access. BMJ. 2023 Feb 3;380:290

101 McMullin J. Cancer. Annu Rev Anthropol. 2016;45:251–66.

102 Zelek L, Sargent C. Réflexions sur la prise en charge des femmes originaires de l’Afrique de l’Ouest en cancérologie. La santé en Action. 2021 March;(455): 34–7.

103 Espina C, McKenzie F, Dos-Santos-Silva I. Delayed presentation and diagnosis of breast cancer in African women: a systematic review. Ann Epidemiol. 2017 Oct;27(10):659–71.

104 Dumalaon-Canaria JA, Hutchinson AD, Prichard I, Wilson C. What causes breast cancer? A systematic review of causal attributions among breast cancer survivors and how these compare to expert-endorsed risk factors. Cancer Causes Control. 2014 Jul;25(7):771–85.

105 Neishaboury M, Davoodzadeh K, Karbakhsh M. does embarrassment contribute to delay in seeking medical care for breast cancer? Arch Breast Cancer. 2015:2(3):75–8.

106 Sargent C, Larchanche S. Transnational health-care circuits: Managing therapy among immigrants in France and kinship networks in West Africa. In: Cole J, C Groes, eds. Affective Circuits. African Migrations to Europe and the Pursuit of Social Regeneration. Chicago: Chicago University Press, 2016, pp. 101–125.

107 Al-Azri M, Al-Awisi H, Al-Moundhri M. Coping with a diagnosis of breast cancer-literature review and implications for developing countries. Breast J. 2009 Nov-Dec;15(6):615–22.

108 Schantz C, Coulibaly A, Faye K, Traoré D; SENOVIE group. Amazons in Mali? Women’s experiences of breast cancer and gender (re)negotiation. Soc Sci Med. 2024 May;348:116874.

109 Estupiñán Fdez de Mesa M, Ferguson M, Green S, Marcu A, Ream E, Whitaker KL. Applying the Intersectionality Lens to Understand Minority Ethnic Women’s Experiences of the Breast Cancer Care Pathway in England: A Qualitative Interview Study. Psychooncology. 2025 Feb;34(2):e70092.

110 Drageset S, Lindstrøm TC. Coping with a possible breast cancer diagnosis: demographic factors and social support. J Adv Nurs. 2005 Aug;51(3):217–26.

111 Al-Azri M, Al-Awisi H, Al-Moundhri M. Coping with a diagnosis of breast cancer-literature review and implications for developing countries. Breast J. 2009 Nov-Dec;15(6):615-22.

112 Pleasant V. A public health emergency. Breast cancer among Black communities in the United States. Obstet Gynecol Clin N Am. 2024;51:69–103.

113 Moodley J, Unger-Saldaña K. The role of racial and ethnic discrimination in breast cancer disparities. Lancet. 2024 May 11;403(10439):1827–1830.

114 Chen T, Kharazmi E, Fallah M. Race and Ethnicity-Adjusted Age Recommendation for Initiating Breast Cancer Screening. JAMA Netw Open. 2023 Apr 3;6(4):e238893.

